# Survival Outcomes after Monitoring, Surgery, or Radiotherapy for Clinically Localized Prostate Cancer

**DOI:** 10.1101/2025.07.16.25331661

**Authors:** Min Ho An, Chungsoo Kim, Kyungchan Min, Kyu-Ho Yi, Rae Woong Park, Minji Jung

## Abstract

**Purpose:** A recent randomized controlled trial, ProtecT, found similar survival benefits between active surveillance (AS), radiotherapy (RT), or radical prostatectomy (PT) in localized prostate cancer (PC), while observational studies showed conflicting results. This study aims to emulate the ProtecT trial to compare survival outcomes in men with localized PC who underwent PT or RT compared to AS/watchful waiting (WW).

**Methods:** This target trial emulation study used the Korean Nationwide Health Insurance, Cancer Registry, and Death Registry linkage data (2012-2020). We included men (50-69 years) diagnosed with a first malignant localized PC. The primary outcome was PC-specific death, and secondary outcomes were all-cause death, metastasis, and antidepressant initiation. Propensity score matching created two balanced comparisons: PT vs AS/WW and RT vs AS/WW. Cox regression was used to calculate hazard ratios (HR) with 95% confidence interval (CI). Two sensitivity analyses were conducted, by including individuals aged over 40 and those with CVD history.

**Results:** Among 8,036 localized PC patients, 1,191 PT vs 1,191 AS/WW and 428 RT vs 428 AS/WW individuals were identified. During a mean follow-up of 4.4 years, PC-specific death occurred in 2 men (0.2%) in PT and 2 men (0.2%) in AS/WW, and 3 men (0.7%) in RT with 1 man (0.2%) in AS/WW. AS/WW showed comparable risks of PC-specific and all-cause death and metastasis as PT and RT. Compared to AS/WW, PT and RT showed lower risks of antidepressant initiation (HR [95% CI] 0.73 [0.58-0.93] for PT and HR 0.38 [0.24-0.61] for RT, respectively). Two sensitivity analyses showed robustness.

**Conclusions:** In this target trial emulation study, AS/WW showed similar risks of mortality and metastasis as PT and RT, while PT and RT showed favorable outcomes with antidepressant initiation in Asian men with localized PC.

## Introduction

Prostate cancer (PC) is one of the most common cancers worldwide, with over 1.4 million new cases annually.^1^ In 2024, it has been the most frequently diagnosed cancer among Korean men, accounting for 15.8% of new male cancers and ranking among the top seven causes of male cancer deaths.^2^ The era of prostate-specific antigen (PSA) testing has increased the diagnosis rates of localized PC,^3^ for which treatment options include prostatectomy (PT), radiotherapy (RT), and active surveillance/watchful waiting (AS/WW).

Although AS/WW is often preferred due to its potential to minimize overtreatment, reduce treatment-related complications, and preserve quality of life,^4,5^ identifying the most effective and safe treatment remains challenging due to the inconsistent results shown by existing randomized controlled trials (RCTs), which are considered the gold standard for treatment evaluation. The ProtecT trial, the most recent trial enrolling men with localized PC diagnosed in the PSA era, found no significant difference in PC-specific and all-cause death between PT, RT, and AS, though PT and RT were more effective in preventing metastases and progression.^6^ The PIVOT trial, conducted during the earlier PSA era, also showed no significant difference between PT and WW.^7^ In contrast, the SPCG-4 trial, conducted in a pre-PSA era population, showed a survival advantage for PT over WW.^8^ Observational studies from the US and Canada also favored PT and RT over AS/WW for better survival outcomes.^9–12^

Traditional observational studies are often limited by biases, such as selection, confounding, or time-related biases. Target trial emulation addresses these limitations by using real-world data to simulate RCTs, reflecting the heterogeneity of routine clinical practice.^13^ This approach enables the evaluation of treatment effects based on emulated RCT protocol across diverse settings, including underrepresented populations such as individuals with comorbidities, older adults, and racial and ethnic minorities, including Asians. Therefore, we aimed to emulate the ProtecT trial by comparing PT and RT with AS/WW in an Asian population.

## Methods

### Data Source

This study utilized the national integrated cancer dataset consisting of the Korean Central Cancer Registry (KCCR) data from 2012 to 2021, national claims from the Korean Nationwide Health Insurance and Screening, and the National Death Registry from the Korean Statistical Information Service, provided through the Korea-Clinical data Utilization network for Research Excellence (K-CURE) platform.^14^ As Korea operates the single-payer national health insurance system and maintains the national cancer registry,^15,16^ the K-CURE platform includes comprehensive data on all registered patients with cancer, the entire cancer population in Korea.^14^ Detailed information of each dataset is described in Table S1. This study was reviewed by the institutional review board (IRB number: AJOUIRB-EX-2024-348) of Ajou University Medical Center.

### Study Design and Study Population

In this target trial emulation study, we followed the protocol of ProtecT trial. We included male patients diagnosed with a first primary, histologically confirmed, clinically localized PC. The localized PC was defined based on records in the cancer registry with PC diagnosis (International Classification of Diseases 10^th^ revision [ICD-10] Code, C61 with descendants) and cancer stage classified as ‘localized’. To minimize potential immortal time bias by identifying “time-zero”, we first defined a cohort entry as the date of PC diagnosis and designated the following four months as the screening period to identify study groups based on their initial treatment. This four-month period aligns with the time frame to classify the first course of treatment in the KCCR data. The index date, the time-zero for follow-up, was then set to be four months (120 days) after cohort entry. Figure S1 shows schematic diagram for the cohort identification and Table S2 provides detailed information on the trial criteria compared to the emulation criteria.

### Study Exposures

Three study groups, AS/WW, RT or PT, were identified based on the first course of treatment during the four months post-PC diagnosis. The AS/WW group was defined as patients who did not receive any definitive treatment (i.e., surgery, radiotherapy, hormonal therapy, or chemotherapy). Additionally, to align with the AS group in the ProtecT, which monitored PSA levels every three months during the first year and every 6 to 12 months thereafter, we applied an additional criterion: patients in the AS/WW group must have undergone at least one confirmatory screening tests, including PSA tests, magnetic resonance imaging, ultrasonography, biopsy, or digital rectal exam, within four months after the diagnosis. However, since we were unable to fully differentiate AS from WW using the administrative claims due to potential immortal time bias, we referred to this group collectively as the AS/WW group. The PT and RT groups were defined as patients with a record of radiotherapy and surgical procedures, respectively, within the four-month period. Detailed codes are listed in Table S2 & S3.

### Study Outcomes

The primary outcome was PC-specific death. Secondary outcomes were all-cause death, metastasis, and antidepression initiation. PC-specific death was determined based on the primary cause of death in the National Death Registry. Metastasis was identified using ICD-10 codes (C77-C79). Antidepressant initiation, which is prevalent among men with PC; approximately 1 in 6 patients experience clinical depression,^17^ was defined as the first prescription record of any antidepressants (Table S3). Patients were followed as an intent-to-treat manner, from the index date until the earliest of the following dates: date of the study outcome, date of death, or end of the observation period (December 31, 2021).

### Covariates

Demographic information were identified at the PC diagnosis, including age, income level (low, middle, and high tertiles), and the year groups of PC diagnosis (2013-2014, 2015-2017, and 2018-2019). Lifestyle factors, including BMI, and smoking status (never, former, and current smoker), were identified from the latest health screening data within two years prior to the cohort entry, considering the biennial cycle of national health screening program. Comorbidities and comedication use were identified during one year prior to the PC diagnosis. Comorbidities included hypertension, dyslipidemia, diabetes mellitus, and benign prostatic hyperplasia (BPH), using ICD-10 codes. Additionally, Charlson Comorbidity Index (CCI) was calculated based on each patient’s medical history.^18^ Comedication use included 5-alpha reductase inhibitors (5-ARI), anticholinergics, alpha-blockers, corticosteroids, and beta 3 adrenoreceptor agonist (B3AR) (Table S3).

### Statistical Analysis

Categorical variables were reported as counts and percentages. Continuous variables were reported as means with standard deviations or as medians and interquartile ranges. We used propensity score (PS) matching for two paired groups: PT vs AS/WW and RT vs AS/WW. PS was calculated by fitting a multivariable logistic regression model incorporating the following covariates: age, income levels, PC diagnosis year groups, CCI, dichotomized comorbidity (BPH) and comedication use (5-ARI, alpha-blockers, anticholinergics, and B3AR), BMI, and smoking status. We utilized the greedy nearest neighbor matching method with a caliper value of 0.01 at a 1:1 ratio. Covariates were considered balanced when a standardized mean difference (SMD) of less than 0.1.^19^ The incidence rate with 95% confidence interval (CI) was calculated as the number of new outcome cases per person-year at risk during the defined study follow-up period. We performed the Cox proportional hazard regressions to estimate hazard ratio (HR) of the study outcomes and its 95% CI. The Fine and Gray competing risk regressions were conducted to evaluate PC-specific death, while accounting for competing risk of deaths from other causes, e.g., cardiovascular death. Cumulative incidence curves were plotted and a log-rank test was performed.

Since age is a strong confounder in the association between treatment modalities and cancer outcomes,^20,21^ we conducted a stratification analysis by age groups (<65 and ≥65 years) to examine whether the association between study groups and outcomes differed by age groups using interaction terms. Additionally, two sensitivity analyses were performed to enhance the generalizability of findings, beyond the limited inclusion and exclusion criteria of RCTs. First, to assess whether the results remain robust across a broader age group, we expanded the study population to include patients older than 40 years. Second, given previous findings that CVD is the leading cause of non-cancer death in PC patients,^22–24^ we included those with a history of CVD, representing 7.3% of our initial eligible study population. Same analytic approach was applied in these population including PS matching, the Cox and Fine-Gray regressions, and the age stratification. Two-sided P <.05 indicates statistical significance. All analyses were performed using SAS Enterprise Guide, version 8.3 (SAS Institute Inc), and R, version 4.22 (R Foundation for Statistical Computing). Analyses were conducted from January 2024 to November 2024. The reporting of the study followed the Strengthening the Reporting of Observational studies in Epidemiology (STROBE) reporting guideline.

## Results

### Baseline Characteristics

The eligible population consisted of 8,036 localized PC men: AS/WW (N=1,208), PT (N=6,360), and RT (N=468). After PS matching, PT (N=1,191) vs AS/WW (N=1,191) and RT (N=428) vs AS/WW (N=428) were identified and both the matched groups were well balanced (all SMD < 0.1). For PT and AS/WW matched groups, the median (Q1-Q3) age was 63 (60-67) and 63 (60-66), respectively. The median BMI was 24 (22-26) kg/m^2^ and approximately 18% were current smokers in both groups. Overall, more than 95% of patients had BPH, approximately 50% had hypertension or dyslipidemia, and over one-fourth had diabetes (Table 1). Two-thirds used alpha-blockers, around 55% used steroids, and 20% used 5-ARI. Similar distributions of baseline characteristics were observed between the RT and AS/WW groups (Table 1). Baseline characteristics in the main analysis before PS matching and two sensitivity analyses before and after PS matching were also presented in Table S4–S8.

**Figure 1.**
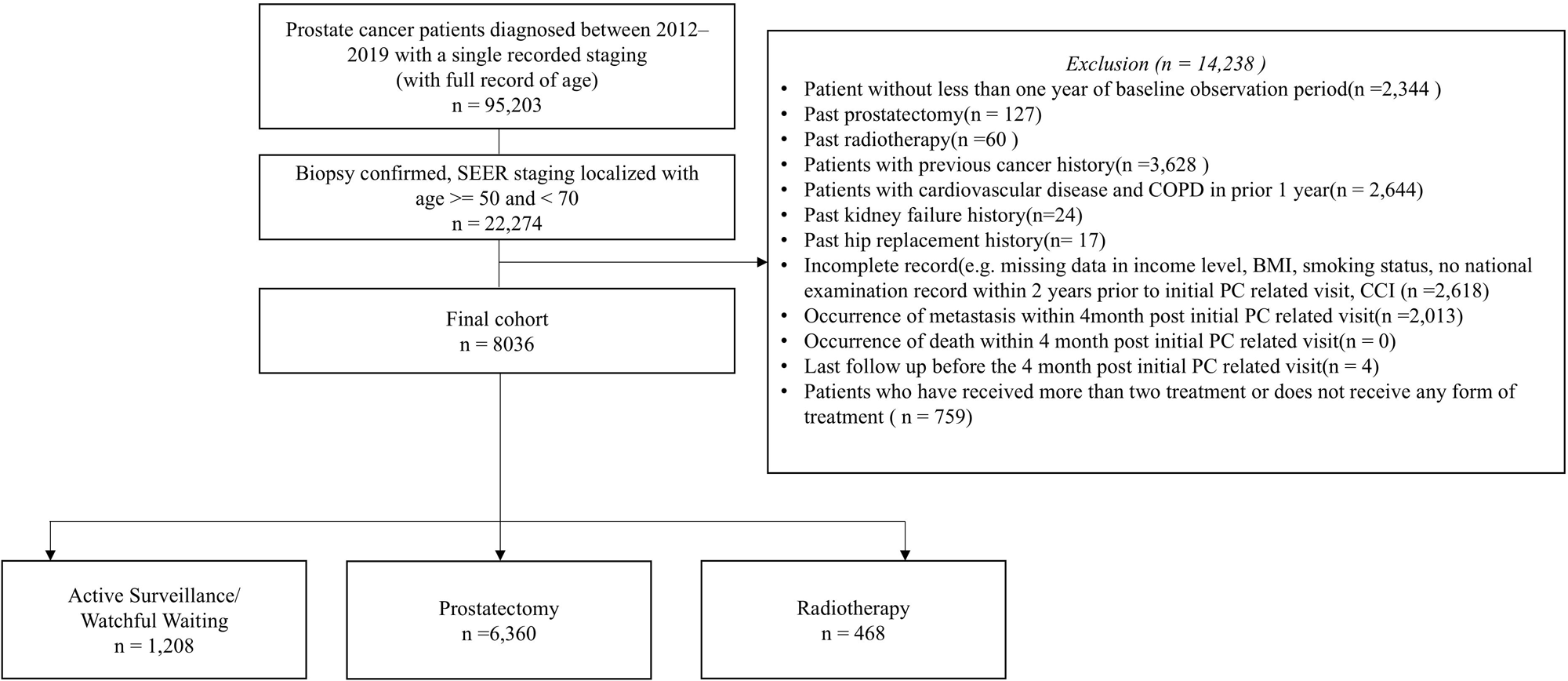
Schematic flow chart of the study population. We included male patients diagnosed with a first primary, histologically confirmed, clinically localized PC. The localized PC was defined based on diagnosis codes with PC (ICD-10 Code, C61 with descendants) and cancer stage classified as ‘localized’. We excluded individuals who met any of the following criteria: 1) non-localized PC diagnosis; 2) age <50 or ≥70 at PC diagnosis; 3) history of any cancer, excluding non-melanoma skin cancer, prior to PC diagnosis; 4) history of cardiovascular disease including stroke, myocardial infarction, and heart failure, and chronic obstructive pulmonary disease within a year preceding PC diagnosis; 5) kidney transplantation or dialysis before PC diagnosis; 6) bilateral hip replacement before PC diagnosis; 7) incomplete one year look-back observation period prior to PC diagnosis; 8) patients who died within the first four-month period (before the index date); and 9) missing or incomplete data on covariates of interests. PC: prostate cancer; ICD-10: International Classification of Diseases, 10th Revision

**Table 1.** Baseline characteristics of the study population after propensity score matching.

| Variables | AS/WW<br>(n = 1,191) | PT<br>(n = 1,191) | SMD | AS/WW<br>(n = 428) | RT<br>(n = 428) | SMD |
| --- | --- | --- | --- | --- | --- | --- |
| Age at PC diagnosis<sup>a</sup> | 63 (60-66) | 63 (60-67) | 0.015 | 63.5 (60-67) | 64 (59-67) | <0.001 |
| PC diagnosis calendar year |  |  | 0.01 |  |  | 0.024 |
| 2013-2014 | 342 (28.7) | 322 (27.0) |  | 76 (17.8) | 78 (18.2) |  |
| 2015-2017 | 233 (19.6) | 226 (19.0) |  | 173 (40.4) | 176 (41.1) |  |
| 2018-2019 | 616 (51.7) | 643 (54.0) |  | 179 (41.8) | 174 (40.7) |  |
| Income levels |  |  | 0.078 |  |  | 0.084 |
| Low | 237 (19.9) | 261 (21.9) |  | 106 (24.8) | 100 (23.4) |  |
| Middle | 256 (21.5) | 278 (23.3) |  | 75 (17.5) | 89 (20.8) |  |
| High | 698 (58.6) | 652 (54.7) |  | 247 (57.7) | 239 (55.8) |  |
| Smoking |  |  | 0.011 |  |  | 0.043 |
| Never | 428 (35.9) | 430 (36.1) |  | 144 (33.6) | 151 (35.3) |  |
| Former | 546 (45.8) | 549 (46.1) |  | 207 (48.4) | 198 (46.3) |  |
| Current | 217 (18.2) | 212 (17.8) |  | 77 (18.0) | 79 (18.5) |  |
| BMI (kg/m<sup>2</sup>)<sup>a</sup> | 24 (22-26) | 24 (22-26) | 0.026 | 24 (23-26) | 24 (23-26) | 0.007 |
| CCI<sup>a</sup> | 4 (2-5) | 4 (2-5) | 0.011 | 4 (2-5) | 4 (2-4.25) | 0.007 |
| Comorbidity |  |  |  |  |  |  |
| Hypertension | 546 (45.8) | 538 (45.2) | 0.013 | 209 (48.8) | 190 (44.4) | 0.089 |
| Dyslipidemia | 590 (49.5) | 586 (49.2) | 0.007 | 223 (52.1) | 214 (50.0) | 0.042 |
| Diabetes mellitus | 322 (27.0) | 326 (27.4) | 0.008 | 128 (29.9) | 114 (26.6) | 0.073 |
| BPH | 1136 (95.4) | 1137 (95.5) | 0.004 | 421 (98.4) | 421 (98.4) | <0.001 |
| Concomitant drug |  |  |  |  |  |  |
| 5-ARI | 224 (18.8) | 244 (20.5) | 0.042 | 92 (21.5) | 86 (20.1) | 0.035 |
| Anticholinergic | 87 (7.3) | 97 (8.1) | 0.031 | 35 (8.2) | 39 (9.1) | 0.033 |
| Alpha-blocker | 789 (66.2) | 777 (65.2) | 0.021 | 302 (70.6) | 298 (69.6) | 0.020 |
| B3AR | 90 (7.6) | 87 (7.3) | 0.010 | 39 (9.1) | 38 (8.9) | 0.008 |
| Steroid | 653 (54.8) | 684 (57.4) | 0.052 | 240 (56.1) | 244 (57.0) | 0.019 |
| Antidepressant | 32 (2.7) | 36 (3.0) | 0.020 | 10 (2.3) | 6 (1.4) | 0.069 |

<sup>a</sup> mean (standard deviation). AS: active surveillance; WW: watchful waiting; RT: radiotherapy; PT: Prostatectomy; SMD: standardized mean difference; SD: standard deviation; PC: prostate cancer; IQR: interquartile range; CCI: Charlson comorbidity index; BPH: benign prostate hyperplasia; 5-ARI: 5-alpha reductase inhibitor; B3AR: beta 3 adrenoreceptor agonist

### Study outcomes

During mean follow-up of 4.4 years, PC-specific death occurred in 2 men (0.2%) in PT and 2 men (0.2%) in AS/WW, and 3 men (0.7%) in RT with 1 man (0.2%) in AS/WW. Comparing PT and AS/WW groups, all-cause death, metastasis, and antidepressant initiation were observed in 12 (1.0%), 117 (9.8%), and 123 (10.3%) men in PT group, versus 15 (1.3%), 117 (9.8%), and 164 (13.8%) men in AS/WW group, respectively. When comparing RT and AS/WW groups, all-cause death, metastasis, and antidepressant initiation occurred in 7 (1.6%), 39 (9.1%), and 24 (5.6%) men in RT group, and 4 (0.9%), 42 (9.8%), and 60 (14.0%) men in AS/WW group, respectively. AS/WW showed similar risks of PC-specific death, all-cause death, and metastasis as PT and RT. Compared to AS/WW, PT and RT showed lower risks of antidepressant initiation (HR 0.73 [0.58-0.93] for PT and HR 0.38 [0.24-0.61] for RT, respectively) (Table 2, Figure 2, and Figure S2 & S3). When stratified by age groups, consistent findings supported the main findings (Table 3).

**Figure 2.**
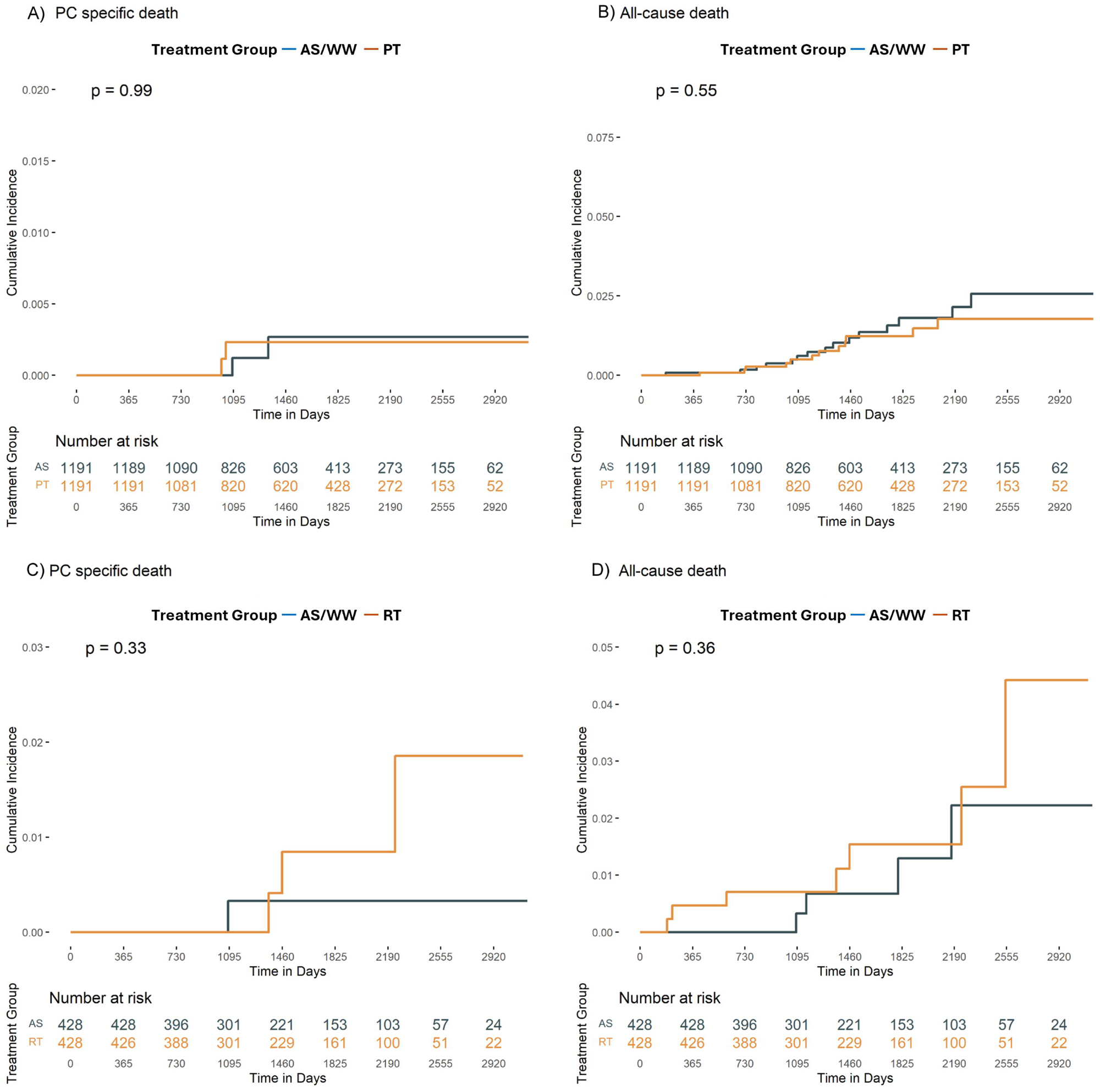
Cumulative incidence plots for prostate cancer specific mortality and all-cause mortality. AS/WW showed similar risks of PC-specific death and all-cause death as PT and RT. AS/WW: active surveillance/watchful waiting; RT: radiotherapy; PT: prostatectomy; HR: hazard ratio; CI: confidence interval

**Table 2.** Incidence and hazard ratios of primary and secondary outcomes.

|  | **AS/WW**<br>**N=1 191** | **PT**<br>**N=1 191** | **AS/WW**<br>**N=428** | **RT**<br>**N=428** |
| --- | --- | --- | --- | --- |
| **Primary outcome** |  |  |  |  |
| **Death from prostate cancer** |  |  |  |  |
| No. of events (%) | 2 (0.2) | 2 (0.2) | 1 (0.2) | 3 (0.7) |
| PY | 5205 | 5222 | 1894 | 1889 |
| IR [95% CI] | 0.38 [0.05-1.39] | 0.38 [0.05-1.38] | 0.53 [0.01-2.94] | 1.59 [0.33-4.64] |
| HR [95% CI] | Reference | 1.01 [0.14-7.16] | Reference | 2.95 [0.31-28.39] |
| **Secondary outcomes** |  |  |  |  |
| **Death from any cause** |  |  |  |  |
| No. of events (%) | 15 (1.3) | 12 (1.0) | 4 (0.9) | 7 (1.6) |
| PY | 5205 | 5222 | 1894 | 1889 |
| IR [95% CI] | 2.88 [1.61-4.75] | 2.30 [1.19-4.01] | 2.11 [0.76-5.41] | 3.71 [1.49-7.64] |
| HR [95% CI] | Reference | 0.80 [0.37-1.70] | Reference | 1.76 [0.52-6.02] |
| **Metastasis** |  |  |  |  |
| No. of events (%) | 117 (9.8) | 117 (9.8) | 42 (9.8) | 39 (9.1) |
| PY | 4866 | 4868 | 1753 | 1804 |
| IR [95% CI] | 24.0 [19.9-28.8] | 24.0 [19.9-28.8] | 24.0 [17.3-32.4] | 21.6 [15.4-29.5] |
| HR [95% CI] | Reference | 0.99 [0.77-1.29] | Reference | 0.90 [0.58-1.40] |
| **Antidepressant initiation** |  |  |  |  |
| No. of events (%) | 164 (13.8) | 123 (10.3) | 60 (14.0) | 24 (5.6) |
| PY | 4721 | 4852 | 1727 | 1815 |
| IR [95% CI] | 34.7 [29.6-40.5] | 25.4 [21.1-30.2] | 34.7 [26.5-44.7] | 13.2 [8.47-19.7] |
| HR [95% CI] | Reference | **0.73 [0.58-0.93]** | Reference | **0.38 [0.24-0.61]** |

AS/WW: active surveillance/watchful waiting; PT: prostatectomy; RT: radiotherapy; No: number; PY: person-year; IR: incidence rate per 1,000 person-year; CI: confidence interval; HR: hazard ratio.

**Table 3.** Results of subgroup analysis by age.

| **Outcomes** | **Events (n) in**<br>**AS/WW / PT** | **PT vs AS/WW**<br>**HR [95% CI]** | **P-value for**<br>**interaction** | **Events (n) in**<br>**AS/WW / RT** | **RT vs AS/WW**<br>**HR [95% CI]** | **P-value for**<br>**interaction** |
| --- | --- | --- | --- | --- | --- | --- |
| **Death from prostate cancer** |  |  | NA |  |  | NA |
| < 65 years | 2/1 | 0.53 [0.05-5.85] |  | 1/1 | 1.11 [0.07-17.78] |  |
| ≥ 65 years | 0/1 | NA |  | 0/2 | NA |  |
| **Death from any cause** |  |  | 0.392 |  |  | 0.694 |
| < 65 years | 9/5 | 0.57 [0.19-1.71] |  | 3/4 | 1.51 [0.34-6.76] |  |
| ≥ 65 years | 6/7 | 1.21 [0.38-3.34] |  | 1/3 | 2.59 [0.27-24.91] |  |
| **Metastasis** |  |  | 0.727 |  |  | 0.806 |
| < 65 years | 68/63 | 0.95 [0.68-1.35] |  | 24/21 | 0.4 [0.52-1.69] |  |
| ≥ 65 years | 49/54 | 1.06 [0.71-1.55] |  | 18/18 | 0.84 [0.44-1.62] |  |
| **Antidepressant initiation** |  |  | 0.807 |  |  | 0.354 |
| < 65 years | 99/70 | **0.71 [0.53-0.97]** |  | 37/16 | **0.46 [0.25-0.82]** |  |
| ≥ 65 years | 65/53 | 0.76 [0.53-1.09] |  | 23/8 | **0.28 [0.12-0.62]** |  |

AS/WW: active surveillance / watchful waiting; PT: prostatectomy; RT: radiotherapy; n: number; PY: person-year; CI: confidence interval; NA: not available.

### Sensitivity Analyses

Two sensitivity analyses showed robust findings (Table S9–S10 and Figure S4–S7). First, expanding the study population to include individuals aged over 40, PT (n=2239) showed similar hazards of PC-specific death, all-cause deaths, metastasis, and antidepressant initiation compared to AS/WW (n=2239). Similarly, RT (n=1160) and AS/WW (n=1160) had comparable risks of PC-specific death, all-cause deaths, and metastasis, while RT was associated with a lower risk of antidepressant initiation (HR 0.69 [0.54-0.89]). Second, when including patients with a history of CVD, PT (n=1332) and RT (n=491) showed comparable risks for all outcomes relative to their respective AS/WW groups, except for RT, which was associated with a lower risk of antidepressant initiation (HR 0.52 [0.35-0.78]). Applying the Fine and Gray regressions, similar results were shown (Table S11).

## Discussion

This is the first target trial emulation, to our knowledge, to compare the effects of treatment modalities among men with PSA-detected, clinically localized PC. By emulating the ProtecT trial, we observed high survival rates (99.8% from PC-specific death and 98.8% from all-cause death) across all treatment groups during a mean follow-up of 4.4 years in an Asian population. Consistent with the trial findings, AS/WW showed similar risks of PC-specific and all-cause deaths as well as metastasis when compared to PT and RT. However, PT and RT were associated with a lower risk of antidepressant initiation compared to AS/WW. These associations were consistent in both younger and older age groups. Additionally, findings from two expanded sensitivity analyses supported the robustness of the main findings.

Our findings align with those of the ProtecT trial, which reported results with a median follow-up of 15 years.^6^ Although our study had a relatively shorter follow-up, resulting in lower cumulative mortality rates, it is novel in demonstrating comparable survival outcomes between active, radical treatments and observation in Asian men with localized PC. The target trial also assessed the risk of clinical progression and the initiation of long-term androgen deprivation therapy (ADT) as additional secondary outcomes. While the trial showed that PT and RT significantly reduced these risks by half compared to AS, these reductions did not translate into differences in survival outcomes.^6^ Taken together, these findings suggest that AS/WW may be a more considered initial treatment option in localized PC men to balance survival benefits against the risk of overtreatment such as urinary incontinence, sexual dysfunction, bowel dysfunction, or lymphedema.^25,26^.

Our main findings differ from previous traditional observational studies, where suggested that PT and RT are associated with survival advantages compared to AS/WW.^9–12,27,28^ We emulated the target trial with stricter eligibility criteria, including age restriction (50-69 years) and exclusion criteria of patients with specific comorbidities such as CVD, and definition of treatment assignment (confirmatory screening test based AS/WW group) compared to traditional observational studies (various age ranging from 30 to 95 years or no such exclusion criteria or definition). In our sensitivity analysis, which expanded the study population to include individuals aged over 40 years, we observed similar results: PT was associated with a decreased risk of PC-specific death compared to AS/WW but had no impact on all-cause death. Older age might influence initial treatment decisions and survival outcomes in men with localized PC. Life expectancy is a key factor in choosing initial treatments.^4^ Despite balanced age distribution and other baseline characteristics between PT and AS/WW groups, unmeasured confounders are inevitable, and older patients may be more likely to opt for less aggressive treatments such as AS/WW, potentially influencing survival outcomes differently. Since CVD-related death is one of the leading causes of death among men with localized PC,^22–24^ a CVD history may confound the associations between initial treatment options and survival outcomes. However, similar results were observed when including individuals with a history of CVD in our sensitivity analysis.

We applied PS matching to reduce confounding and selection bias and corresponding statistical analysis to the target trial. Moreover, we established a precise and well-defined follow-up starting point (time zero) to effectively mitigate immortal time bias, which occurs when treatment is assigned after follow-up begins. In contrast, previous studies often lacked a clearly specified starting point^10–12^ or often defined it as PC diagnosis date.^27^ Among the limited exception, one study conducted a 2-year landmark analysis,^9^ which inherently restricted its population to individuals who survived at least two years after diagnosis, resulting in a fundamentally different cohort from ours.

Beyond these, several factors may also explain the different findings,^9–12,27^ including variations in healthcare system. Differences in clinical practices and access disparities could be critical factors. Compared to the US, the single-payer, public healthcare system in Korea ensures widespread access to screening, diagnosis, and treatment, potentially leading to earlier detection, uniform care quality, and greater accessibility. This might help mitigate survival differences between AS/WW and definitive treatments. Variances in study designs may also play a role, such as variations in period of study (1990s-2010s), which correspond to the PSA era and evolving diagnostic and therapeutic approaches, statistical methods (PS matching, weighting, or none) and median follow-up period (ranging from 3 to 12 years).

Interestingly, our emulation study found that PT and RT were associated with a lower risk of antidepressant initiation compared to AS/WW. Uncertainty of living with untreated cancer may cause depression or related symptoms, such as anxiety or distress, in patients with PC,^29^ while the close monitoring required for PC patients on AS may further exacerbate these concerns.^30^ Moreover, moderate or severe urinary symptoms in men with localized PC on AS may increase the related concerns.^31^ However, prior research found no differences in depression or anxiety between PT and AS/WW in men with localized PC.^32–34^ Patient-reported outcomes from the ProtecT trial showed no differences up to five years post-diagnosis.^32^ Different methods for measuring depression could contribute to varying results; prescribing antidepressants typically reflects more severe symptoms compared to questionnaire-based assessments used in most of the previous studies.^32–34^ The associations between depression risk and treatment options in patients with localized PC remains controversial, highlighting the need for further studies to validate and expand upon our findings.

Our emulation study demonstrated consistent findings regarding the comparable effects of observation and active treatment in Asian men with localized prostate cancer, aligning with the target trial. However, these findings should be interpreted with caution. First, although we explicitly emulated the trial, disease progression and initiation of long-term ADT were not assessed due to unclear definitions (e.g., type of therapy and duration) in the protocol and potential misclassification. Second, due to the limited information on Gleason scores and PSA levels, which are critical for risk stratification for treatment decision,^4,35^ as well as patient preferences and life expectancy at diagnosis, which can also influence treatment choices, unmeasured or residual confounding is inevitable. Third, the relatively short follow-up period limited our data to assess long-term survival and perform further subgroup analyses due to the low cumulative number of events. Fourth, the high prevalence of BPH in our study may partly be due to BPH often being included in the differential diagnosis to prescribe alpha-blockers in such cases. However, since both groups were well-balanced, this suggests potential non-differential misclassification bias, likely biasing the results toward the null.

## Conclusions

In this target trial emulation study, AS/WW showed comparable effects of PC-death, all-cause death, and metastasis compared to PT and RT in Asian men with localized PC. PT and RT showed a significantly lower risk of antidepressant initiation compared to AS/WW.

## Data availability

This study utilized the national integrated cancer dataset consisting of the Korean Central Cancer Registry data, the Korean Nationwide Health Insurance and Screening data, and the Korean Statistical Information Service data, provided through the Korea-Clinical data Utilization network for Research Excellence (K-CURE) platform. The authors cannot legally distribute the data, but details on data can be found here: https://k-cure.mohw.go.kr/portal/cmm/htm/viewEngMain.do.

## Author Contributions

MA: Data curation, Methodology, Investigation, Formal analysis, Visualization, Writing – review & editing, Funding. CK: Methodology, Investigation, Validation, Writing – review & editing. KM: Writing – review & editing. KY: Writing – review & editing. RP: Supervision, Methodology, Conceptualization, Writing – review & editing, Funding. MJ: Supervision, Methodology, Conceptualization, Writing – original draft & review.

