## Supplementary file for "Survival Outcomes after Monitoring, Surgery, or Radiotherapy for Clinically Localized Prostate Cancer"

**Supplementary Material**

MH An, C Kim et al., Supplementary material to “**Survival Outcomes after Monitoring, Surgery, or Radiotherapy for Clinically Localized Prostate Cancer: Target Trial Emulation**”

### **Table S1. Description of data sources from the Korea Clinical data Utilization network for Research Excellence (K-CURE) platform**

| **Name** | **Description** |
| --- | --- |
| **Korean Central Cancer Registry (KCCR)** | **General description:** This study used data from the National Cancer Registration Program, a nationally collected clinical registry for patients with cancer, covering the period from 2012 to 2019.  **Data elements:** Age, sex, cancer diagnosis using ICD-10 codes, topography (benign, carcinoma in situ, primary or secondary malignancy, and metastasis), first diagnosed date of cancer, Surveillance, Epidemiology, and End Results cancer stage (localized, regional, distant, and unknown), and type of first treatment course within four months following the initial diagnosis (surgery, chemotherapy, radiotherapy, immunotherapy, hormonal therapy, none/unknown, or combined). |
| **Korean Nationwide Health Insurance and Screening (KNHIS)** | **General description:** This study used data from the national health insurance claims and health screening database (2012-2021), which operates under Korea’s mandatory single-payer health insurance system that covers 98% of Korean population.  **Data elements:** Age, sex, income level, diagnosis, procedure, and prescription records from both outpatient and inpatient settings, and lifestyle factors collected from biennial national health checkup/survey including body mass index (BMI), and smoking status. |
| **National Death Registry by Korean Statistical Information Services (KSIS)** | **General description:** This study utilized death data from the national death registry, managed by the Korean statistical information service, that records all deaths occurring in Korea, covering the period from 2012 to 2019.  **Data elements:** Death date and cause of death (primary and secondary coded with ICD-10). |

ICD: International Classification of Diseases

### **Table S2. Specification of target trial emulation for evaluating the effects of different treatment strategies in localized prostate cancer**

| Component | Target Trial | Emulated trial using clinical data |
| --- | --- | --- |
| Aim | To evaluate the effectiveness of three treatments (active monitoring, radical prostatectomy and radical radiotherapy) for men with localized prostate cancer. | To evaluate the effectiveness of three treatments (active monitoring/watchful waiting, radical prostatectomy and radical radiotherapy) for men with localized prostate cancer. |
| Eligibility criteria | Inclusion criteria   - Age 50-69 years on the data of the prostate check clinic - Male gender - Able to give informed written consent to participate - Fit for any of the three treatments   Exclusion criteria – Participants will be excluded from entry if they have   - Concomitant or past malignancies (other than a small, treated skin cancer) - Prior treatment for prostate malignancy - Serious cardiac problems in the previous 12 months of the prostate check clinic, i.e., stroke, myocardial infarction, heart failure - Kidney dialysis or transplantation - Bilateral hip replacement | Inclusion criteria   - Same, except that receiving the informed consents and fit for any of the three treatments did not need to be considered.   Exclusion criteria   - Same - Note: individuals with chronic obstructive pulmonary disease, kidney dialysis, bilateral hip replacement which was identified within the prior 1 year of prostate cancer diagnosis. |
| Treatment strategy | Receiving active surveillance   - Patient were measured on prostate-specific antigen levels every 3 months during the first year of the trial and every 6 to 12 months   Receiving prostatectomy   - In the prostatectomy group, the use of adjuvant of salvage radiotherapy was discussed with patients who had positive surgical margins, extracapsular disease, or a postoperative PSA level of 0.2 ng/mL or higher   Receiving radiotherapy   - Radiotherapy was delivered along with neoadjuvant androgen-deprivation therapy for 3 to 6 months with three-dimensional conformal radiotherapy at 74 Gy in 37 fractions | Receiving active surveillance/watchful waiting   - Patients who did not receive any definitive treatment (i.e., surgery, radiotherapy, hormonal therapy, or chemotherapy) and who underwent at least one confirmatory screening tests, including PSA tests, magnetic resonance imaging, ultrasonography, biopsy, or digital rectal exam, within four months after prostate cancer diagnosis.   Receiving prostatectomy   - Patients who had a record of surgical procedures within four months after prostate cancer diagnosis.   Receiving radiotherapy   - Patients who had a record of radiotherapy within four months after prostate cancer diagnosis. |
| Treatment assignment | Eligible patients are randomly assigned to three treatment groups. | To generate a study population with similar probability of treatment randomized assignments among the three treatments, we applied propensity score matching approaches and created two separate cohorts:   - Prostatectomy vs active surveillance/watchful waiting - Radiotherapy vs active surveillance/watchful waiting.   To emulate the random treatment assignment, a propensity score matching was applied. Propensity score was calculated by fitting a multivariable logistic regression model incorporating the following covariates:   - Continuous: age, Charlson Comorbidity Index, body mass index - Categorical: income level, prostate cancer diagnosis year group, smoking status, comorbidity (benign prostatic hyperplasia), concomitant medication (5-alpha reductase inhibitor, alpha-blocker, anticholinergics, and beta-3 adrenoreceptor agonist).   We applied the greedy nearest neighbor matching with a caliper value of 0.01 at a 1:1 ratio. |
| Follow-up | Follow-up begins at treatment assignment and ends at occurrence of outcome. Censored at the time that the men were lost to follow-up. | Patients were followed from four months after initial prostate cancer diagnosis until the earliest of the following dates: date of outcome occurred, date of death, or end of the observation period (December 31, 2021). The four-month period was defined as a treatment randomized assignment window. |
| Outcomes | Primary outcome   - The primary outcome is definite or probable prostate cancer specific mortality (including definite or probable intervention-related mortality) at a median of 10-, 15-, and 20-years following randomization.   Secondary outcomes   - Overall survival at a median of 10-, 15-, and 20-years follow-up. - Other clinical outcomes (disease progression, treatment complications) at up to 10-, 15-, and 20-years follow-up. - Urinary and bowel symptoms quality of life, sexual function, depression and other psychosocial effects up to 15-years follow-up. | Primary outcome   - Same, except follow-up period in our data is a maximum 9 years (Jan 2013- Dec 2021).   Secondary outcomes   - Overall survival - Metastasis - Initiation of antidepressants |
| Causal contrast | Intention-to-treat framework   - Included in the analysis for as long as they were undergoing clinical follow-up | Same |
| Statistical analysis | Cox proportional hazard regression after adjustment for trial center, patient’s age, Gleason score, and baseline PSA (log-transformed) to compare prostate cancer specific mortality at 15 years in the three groups on an intention-to-treat basis.  Pairwise significance tests were planned if the p-value for equal disease-specific mortality across the trial groups was less than 0.05 (on the basis of an overall false positive risk of 5%). Interaction terms were added to this model to investigate differential treatment effects across the eight prespecified subgroups. | After applying the propensity score matching, univariable Cox proportional hazards regression was used to estimate hazard ratios of the study outcome in the three group on an intention-to-treat basis.  The Fine and Gray competing risk regression was conducted to evaluate prostate cancer specific mortality, while accounting for competing risk of deaths from other causes, e.g., cardiovascular death. |

### **Table S3. Code lists to define study population and outcomes**

| **Domain** | **Category** | **Variable** | **Code** |
| --- | --- | --- | --- |
| Diagnosis  (ICD-10) | Outcomes, Inclusion/Exclusion criteria, comorbidities | Malignant tumor (except for mild skin cancer) | C00-C97, except for C43-C44 |
|  |  | Stroke | I60-I64, G45 |
|  |  | Myocardial infarction | I21-I22, I24, I25 |
|  |  | Heart failure | I50 |
|  |  | Chronic obstructive pulmonary disease | J46-J44 |
|  |  | Secondary neoplasm or metastasis | C76-80, C81-96, C97 |
|  |  | Hypertension | I10-15% |
|  |  | Dyslipidemia | E78% |
|  |  | Diabetes mellitus | E10-14% |
|  |  | Benign prostate hyperplasia | N40% |
| Procedures (Korean EDI code) | Prostatectomy | Prostatectomy | R3950 |
|  |  | Total Prostatoseminal Vesiculectomy | R3960 |
|  |  | Cryosurgical Ablation of Prostate Cancer | RZ512 |
|  | Radiotherapy | Conformal radiation therapy | HD061 |
|  |  | 3-Dimensional Conformal Radiotherapy (≥2.5 Gy, <5.0 Gy) | HD463 |
|  |  | 3-Dimensional Conformal Radiotherapy (≥ 5.0 Gy) | HD464 |
|  |  | Fractionated stereotactic radiation therapy | HD110 |
|  |  | Stereotactic radiosurgery | HD111 |
|  |  | Body Stereotactic Radiosurgery-LINAC | HD112 |
|  |  | Body Stereotactic Radiosurgery-Cyber Knife | HD211 |
|  |  | Body Stereotactic Radiosurgery(One Time)-Cyber Knife | HD212 |
|  |  | Intensity modulated radiation therapy | HZ271 |
|  |  | Mold therapy | HD080 |
|  |  | Iodine-125 Permanent Implant for Prostate Cancer[Plan] | HD040 |
|  |  | Iodine-125 Permanent Implant for Prostate Cancer[Therapy] | HD150 |
|  |  | Iodine-125 permanent implant for prostate cancer | HZ276 |
|  | PSA measure | Prostate cancer (immunoassay) | D4300 |
|  |  | p2PSA | CZ292 |
|  |  | Prostate cancer (immunoassay) – Nuclear medicine | D4307 |
|  |  | Free PSA | CX321 |
|  |  | Immunoassay: Tumor Marker Test - PSA | B5490 |
|  |  | Prostate Specific Antigen | C4280 |
|  |  | Prostate Specific Antigen – Nuclear medicine | C7428 |
|  |  | Free PSA – Nuclear Medicine | CX732 |
|  | Dialysis | Arterio-Venous Shunt or Fistula Formation for Hemodialysis - External AV Shunt | O2011 |
|  |  | Arterio-Venous Shunt or Fistula Formation for Hemodialysis - Internal AV Shunt | O2012 |
|  |  | Arterio-Venous Shunt or Fistula Formation for Hemodialysis - Fistula Formation：Autologous Vein | O2081 |
|  |  | Arterio-Venous Shunt or Fistula Formation for Hemodialysis - Fistula Formation：Artificial Vein | O2082 |
|  |  | Repair of Arterio-Venous Fistula for Hemodialysis | O2083 |
|  |  | Hemodialysis Access Creation using Com-bination of Arteriovenous Graft and Central Venous Catheter | O2084 |
|  |  | Continuous Venovenous Hemodiafiltration | O7001 |
|  |  | Continuous Venovenous Hemodiafiltration – from the next day | O7002 |
|  |  | Continuous Arteriovenous Hemodiafiltration | O7003 |
|  |  | Continuous Arteriovenous Hemodiafiltration – from the next day | O7004 |
|  |  | Hemodialysis | O7020 |
|  |  | Continuous Venovenous Hemodialysis | O7031 |
|  |  | Continuous Venovenous Hemodialysis – from the next day | O7032 |
|  |  | Continuous Arteriovenous Hemodialysis | O7033 |
|  |  | Continuous Arteriovenous Hemodialysis – from the next day | O7034 |
|  |  | Acute Peritoneal Dialysis - Catheter Insertion | O7061 |
|  |  | Acute Peritoneal Dialysis – Eialysate Exchange | O7062 |
|  |  | Extracorporeal Ascites Dialysis | O7080 |
|  |  | Hemodialysis (Hospital) | O9992 |
|  |  | Hemodialysis (Clinic) | O9993 |
|  | Kidney transplantation | Renal Transplantation | R3280 |
|  | Hip replacement | Replacement Arthroplasty – Total Arthroplasty - Hip | N0711 |
|  |  | Replacement Arthroplasty – Total Arthroplasty – Hip (complex case) | N2070 |
|  |  | Revision of Replacement Arthroplasty – Total Arthroplasty - Hip | N1711 |
|  |  | Revision of Replacement Arthroplasty – Total Arthroplasty – Hip (with removal simultaneously) | N1721 |
|  |  | Revision of Replacement Arthroplasty – Total Arthroplasty – Hip (Complex case) | N3720 |
|  | Biopsy | Prostatic Biopsy - Percutaneous | C8551 |
|  |  | Prostatic Biopsy - Operative | C8552 |
|  | Bone scan | Bone scan – Partial | HC190 |
|  |  | Bone scan – Whole Body | HC191 |
|  |  | Bone scan – Three Phase | HC192 |
|  |  | Bone scan – Pin Hole | HC193 |
|  | Sonography | Male Genital Ultrasound – Prostate Seminal Vesicle | E9447 |
|  |  | Male Genital Ultrasound – Prostate Seminal Vesicle (Transrectum) | EB451 |
|  |  | Male Genital Ultrasound – Prostate Seminal Vesicle (Transabdomen) | EB452 |
|  |  | Abdominal Ultrasound – Rectum | E9445 |
|  |  | Abdominal Ultrasound – Rectum Anus | [EB446](https://opendata.hira.or.kr/op/opc/olapDiagBhvInfoTab1.do#none) |
|  | DRE | Rectal Digital Examination | E7050 |
|  | Orchiectomy | Orchiectomy | R3850 |
|  |  | Orchiectomy – Total | R3851 |
|  |  | Orchiectomy – Undescended Testis | R3852 |
|  |  | Orchiectomy – Partial | R3853 |
|  |  | Orchiectomy for Malignant Tumor | R3860 |
|  |  | Orchiectomy for Malignant Tumor – Including Lymphadenectomy | R3861 |
|  |  | Orchiectomy for Malignant Tumor - Others | R3862 |
|  | MRI | Magnetic Resonance Imaging - Prostate | HE134 |
|  |  | Magnetic Resonance Imaging – Contrast | HE234 |
|  |  | Magnetic Resonance Imaging | HE334 |
|  |  | Magnetic Resonance Imaging – Prostate Limited MRI | HE434 |
|  |  | Magnetic Resonance Imaging – 3D | HE534 |
| Drugs (Korea Drug Code, first five code only) | 5-ARI | dutasteride | 45880 |
|  |  | finasteride | 15900 |
|  | a-Blocker | alfuzosin | 10480 |
|  |  | doxazosin | 14910 |
|  |  | naftopidill | 61420 |
|  |  | phenoxybenzamine | 48340 |
|  |  | phentolamine | 44143 |
|  |  | silodosin | 50420 |
|  |  | tamsulosin | 23460 |
|  | Anticholinergics | fesoterodine | 50380 |
|  |  | flavoxate | 15920 |
|  |  | oxybutynin | 20700 |
|  |  | propiverine | 21970 |
|  |  | solifenacin | 49380, 65560,65740 |
|  |  | tolterodine | 35710 |
|  |  | trospium | 24550 |
|  | Antidepressant | bupropion hydrochloride | 42810 |
|  |  | citalopram hydrobromide | 42830 |
|  |  | desvenlafaxine | 68770 |
|  |  | desvenlafaxine benzoate | 68760 |
|  |  | desvenlafaxine succinate monohydrate | 62640 |
|  |  | dexvenlafaxine succinate | 62640 |
|  |  | duloxetine hydrochloride | 49550 |
|  |  | escitalopram | 52110 |
|  |  | escitalopram oxalate | 47480 |
|  |  | fluoxetine hydrochloride | 16150 |
|  |  | fluvoxamine maleate | 16250 |
|  |  | milnacipran hydrochloride | 35580 |
|  |  | paroxetine hydrochloride | 20930 |
|  |  | venlafaxine hydrochloride | 24750 |
|  | b3 agonist | mirabegron | 62570 |
|  | Hormonal therapy | abiraterone | 62040 |
|  |  | bicalutamide | 11720 |
|  |  | cyproterone acetate | 13940, 39840 |
|  |  | degarelix | 62440 |
|  |  | enzalutamide | 62740 |
|  |  | estradiol hemihydrate | 15490, 29700, 29710, 29740, 43370, 43380, 15493 |
|  |  | estradiol valerate | 15500, 29760, 39840, 43390, 43400, 43410, 50760 |
|  |  | goserelin acetate | 16720 |
|  |  | luprolide acetate | 18263, 18260, 18261 |
|  |  | triptorelin acetate | 24493, 24490 |
|  |  | triptorelin pamoate | 46750 |
|  | Male Hormone | testosterone cypionate | 23613 |
|  |  | testosterone undecanoate | 23630 |
|  | Steroid | betamethasone | 11640, 29690 |
|  |  | betamethasone |  |
|  |  | betamethasone dipropionate | 54710, 54740, 54700, 54730, 54860, 54930, 54870, 54880, 54890, 54900, 54920, 54940, 54970, 54980, 80040, 80050, 80060, 80070, 65750 |
|  |  | betamethasone sodium phosphate | 11653 |
|  |  | betamethasone valerate | 11663, 52830, 52840, 52850, 52860, 52870 |
|  |  | dexamethasone | 14190, 14193, 53790, 53810, 53820, 53980, 53990, 54000, 54010, 54020, 64030, 64300, 64390, 64700, 64710, 54230 |
|  |  | dexamethasone cipecilate | 14190, 14191 |
|  |  | dexamethasone disodium phosphate | 14223 |
|  |  | dexamethasone palmitate | 14203 |
|  |  | dexamethasone propionate | 14213 |
|  |  | dexamethasone sodium phosphate | 14223 |
|  |  | fludrocortisone acetate | 1602 |
|  |  | hydrocortisone | 17090, 54200, 54210, 54220, 17093, 17094, 54660, 54670, 54680, 54690, 17095, 65850, 68590 |
|  |  | hydrocortisone acetate | 17103 |
|  |  | hydrocortisone butyrate | 17113 |
|  |  | hydrocortisone sodium succinate | 17120 |
|  |  | hydrocortisone valerate | 17133 |
|  |  | methylprednisolone | 19330 |
|  |  | methylprednisolone aceponate | 19343 |
|  |  | methylprednisolone acetate | 19353 |
|  |  | methylprednisolone sodium succinate | 19360 |
|  |  | prednisolone | 21703, 21700 |
|  |  | prednisolone acetate | 21713 |
|  |  | prednisolone sodium succinate | 21730 |
|  |  | prednisolone valeroacetate | 21753 |
|  |  | triamcinolone | 24320 |
|  |  | triamcinolone acetonide | 24333, 24334, 54540, 54550, 54560, 54570, 54580, 54590, 80120 |

### **Table S4. Baseline characteristics of study population before the propensity score matching**

| **Variables** | **AS/WW**  **(n = 1,208)** | **PT**  **(n = 6,360)** | **RT**  **(n = 468)** | **SMD** |
| --- | --- | --- | --- | --- |
| Age at PC diagnosis, mean (SD) | 62.6 (4.7) | 62.6 (4.7) | 63.0 (4.8) | 0.054 |
| Income levels |  |  |  |  |
| Low | 239 (19.8) | 1,216 (19.1) | 126 (26.9) | 0.124 |
| Middle | 262 (21.7) | 1,399 (22.0) | 91 (19.4) |  |
| High | 707 (58.5) | 3,745 (58.9) | 251 (53.6) |  |
| PC diagnosis calendar year |  |  |  |  |
| 2013-2014 | 197 (16.3) | 1,563 (24.6) | 99 (21.2) | 0.154 |
| 2015-2017 | 502 (41.6) | 2,638 (41.5) | 188 (40.2) |  |
| 2018-2019 | 509 (42.1) | 2,159 (33.9) | 181 (28.7) |  |
| Smoking |  |  |  |  |
| Never | 430 (35.6) | 2,314 (36.4) | 162 (34.6) | 0.057 |
| Former | 553 (45.8) | 3,041 (47.8) | 220 (47.0) |  |
| Current | 225 (18.6) | 1,005 (15.8) | 86 (18.4) |  |
| BMI (kg/m^2^), mean (SD) | 24.0 (2.5) | 24.0 (2.5) | 24.4 (2.5) | 0.100 |
| CCI, mean (SD) | 3.6 (1.4) | 3.7 (1.3) | 3.7 (1.4) | 0.048 |
| Comorbidity |  |  |  |  |
| Hypertension | 553 (45.8) | 2,854 (44.9) | 215 (45.9) | 0.014 |
| Dyslipidemia | 596 (49.3) | 3,112 (48.9) | 235 (50.2) | 0.017 |
| Diabetes mellitus | 324 (26.8) | 1,703 (26.8) | 127 (27.1) | 0.005 |
| BPH | 1,138 (94.2) | 6,152 (96.7) | 457 (97.6) | 0.117 |
| Concomitant drug |  |  |  |  |
| 5-ARI | 224 (18.5) | 1,370 (21.5) | 99 (21.2) | 0.050 |
| Anticholinergics | 88 (7.3) | 558 (8.8) | 40 (8.5) | 0.037 |
| Alpha-blocker | 792 (65.6) | 4,477 (70.4) | 328 (70.1) | 0.069 |
| B3AR | 91 (7.5) | 411 (6.5) | 44 (9.4) | 0.073 |
| Steroid | 660 (54.6) | 3,561 (56.0) | 265 (56.6) | 0.027 |
| Antidepressant | 32 (2.6) | 156 (2.5) | 8 (1.7) | 0.043 |
| AS: active surveillance; WW: watchful waiting; RT: radiotherapy; PT: Prostatectomy; SMD: standardized mean difference; SD: standard deviation; PC: prostate cancer; IQR: interquartile range; CCI: Charlson comorbidity index; BPH: benign prostate hyperplasia; 5-ARI: 5-alpha reductase inhibitor; B3AR: beta 3 adrenoreceptor agonist. | | | | |

### **Table S5. Baseline characteristics before the propensity score matching in the population in the sensitivity analysis I, which includes patients older than 40 years old**

| **Variables** | **AS/WW**  **(n = 2,263)** | **PT**  **(n = 10,067)** | **RT**  **(n = 1,302)** | **SMD** |
| --- | --- | --- | --- | --- |
| Age at PC diagnosis (mean (SD)) | 68.0 (7.9) | 66.4 (7.2) | 71.3 (8.0) | 0.420 |
| Income levels |  |  |  | 0.055 |
| Low | 438 (19.4) | 1880 (18.7) | 264 (20.3) |  |
| Middle | 418 (18.5) | 1970 (19.6) | 215 (16.5) |  |
| High | 1407 (62.2) | 6217 (61.8) | 823 (63.2) |  |
| PC diagnosis calendar year |  |  |  | 0.149 |
| 2012-2013 | 371 (16.4) | 2459 (24.4) | 272 (20.9) |  |
| 2014-2016 | 958 (42.3) | 4132 (41.0) | 511 (39.2) |  |
| 2017-2019 | 934 (41.3) | 3476 (34.5) | 519 (39.9) |  |
| Smoking |  |  |  | 0.052 |
| Never | 903 (39.9) | 4090 (40.6) | 560 (43.0) |  |
| Former | 1022 (45.2) | 4584 (45.5) | 574 (44.1) |  |
| Current | 338 (14.9) | 1393 (13.8) | 168 (12.9) |  |
| BMI (kg/m^2^), median (IQR) | 23.8 (2.5) | 23.9 (2.5) | 23.9 (2.5) | 0.025 |
| CCI, mean (SD) | 3.7 (1.4) | 3.8 (1.4) | 3.8 (1.4) | 0.058 |
| Comorbidity |  |  |  |  |
| Hypertension | 1189 (52.5) | 4990 (49.6) | 741 (56.9) | 0.098 |
| Dyslipidemia | 678 (30.0) | 3495 (34.7) | 273 (21.0) | 0.207 |
| Diabetes mellitus | 698 (30.8) | 2975 (29.6) | 429 (32.9) | 0.049 |
| BPH | 2175 (96.1) | 9788 (97.2) | 1275 (97.9) | 0.072 |
| Concomitant drug |  |  |  |  |
| 5-ARI | 543 (24.0) | 2530 (25.1) | 391 (30.0) | 0.091 |
| Anticholinergic | 224 (9.9) | 1088 (10.8) | 146 (11.2) | 0.029 |
| Alpha-blocker | 1612 (71.2) | 7370 (73.2) | 1007 (77.3) | 0.093 |
| B3AR | 223 (9.9) | 751 (7.5) | 141 (10.8) | 0.078 |
| Steroid | 1289 (57.0) | 5768 (57.3) | 755 (58.0) | 0.014 |
| Antidepressant | 75 (3.3) | 300 (3.0) | 46 (3.5) | 0.021 |
| AS: active surveillance; WW: watchful waiting; RT: radiotherapy; PT: Prostatectomy; SMD: standardized mean difference; SD: standard deviation; PC: prostate cancer; IQR: interquartile range; CCI: Charlson comorbidity index; BPH: benign prostate hyperplasia; 5-ARI: 5-alpha reductase inhibitor; B3AR: beta 3 adrenoreceptor agonist. | | | | |

### **Table S6. Baseline characteristics after the propensity score matching in the population in the sensitivity analysis I, which includes patients older than 40 years old**

| **Variables** | **AS/WW**  **(n = 2,239)** | **PT**  **(n = 2,239)** | **SMD** | **AS/WW**  **(n = 1,160)** | **RT**  **(n = 1,160)** | **SMD** |
| --- | --- | --- | --- | --- | --- | --- |
| Age at PC diagnosis (mean (SD)) | 67.8 (7.8) | 67.8 (7.1) | 0.004 | 70.4 (7.6) | 70.6 (7.8) | 0.025 |
| Income levels |  |  | 0.017 |  |  | 0.038 |
| Low | 432 (19.3) | 446 (19.9) |  | 239 (20.6) | 239 (20.6) |  |
| Middle | 416 (18.6) | 407 (18.2) |  | 187 (16.1) | 203 (17.5) |  |
| High | 1391 (62.1) | 1386 (61.9) |  | 734 (63.3) | 718 (61.9) |  |
| PC diagnosis calendar year |  |  | 0.017 |  |  | 0.043 |
| 2012-2013 | 370 (16.5) | 384 (17.2) |  | 229 (19.7) | 211 (18.2) |  |
| 2014-2016 | 952 (42.5) | 944 (42.2) |  | 480 (41.4) | 481 (41.5) |  |
| 2017-2019 | 917 (41.0) | 911 (40.7) |  | 451 (38.9) | 468 (40.3) |  |
| Smoking |  |  | 0.059 |  |  | 0.005 |
| Never | 894 (39.9) | 941 (42.0) |  | 483 (41.6) | 483 (41.6) |  |
| Former | 1013 (45.2) | 947 (42.3) |  | 527 (45.4) | 525 (45.3) |  |
| Current | 332 (14.8) | 351 (15.7) |  | 150 (12.9) | 152 (13.1) |  |
| BMI (kg/m^2^), mean (SD) | 23.8 (2.5) | 23.9 (2.5) | 0.014 | 23.9 (2.6) | 23.8 (2.5) | 0.010 |
| CCI, mean (SD) | 3.7 (1.4) | 3.7 (1.4) | 0.014 | 3.8 (1.4) | 3.8 (1.4) | 0.012 |
| Comorbidity |  |  |  |  |  |  |
| Hypertension | 1174 (52.4) | 1159 (51.8) | 0.013 | 653 (56.3) | 647 (55.8) | 0.010 |
| Dyslipidemia | 677 (30.2) | 691 (30.9) | 0.014 | 273 (23.5) | 268 (23.1) | 0.010 |
| Diabetes mellitus | 689 (30.8) | 691 (30.9) | 0.002 | 391 (33.7) | 375 (32.3) | 0.029 |
| BPH | 2158 (96.4) | 2157 (96.3) | 0.002 | 1140 (98.3) | 1135 (97.8) | 0.031 |
| Concomitant drug |  |  |  |  |  |  |
| 5-ARI | 540 (24.1) | 543 (24.3) | 0.003 | 333 (28.7) | 324 (27.9) | 0.017 |
| Anticholinergic | 223 (10.0) | 241 (10.8) | 0.026 | 124 (10.7) | 131 (11.3) | 0.019 |
| Alpha-blocker | 1600 (71.5) | 1576 (70.4) | 0.024 | 880 (75.9) | 883 (76.1) | 0.006 |
| B3AR | 213 (9.5) | 217 (9.7) | 0.006 | 136 (11.7) | 126 (10.9) | 0.027 |
| Steroid | 1275 (56.9) | 1289 (57.6) | 0.013 | 666 (57.4) | 671 (57.8) | 0.009 |
| Antidepressant | 72 (3.2) | 72 (3.2) | <0.001 | 42 (3.6) | 42 (3.6) | <0.001 |
| AS: active surveillance; WW: watchful waiting; RT: radiotherapy; PT: Prostatectomy; SMD: standardized mean difference; SD: standard deviation; PC: prostate cancer; IQR: interquartile range; CCI: Charlson comorbidity index; BPH: benign prostate hyperplasia; 5-ARI: 5-alpha reductase inhibitor; B3AR: beta 3 adrenoreceptor agonist. | | | | | | |

### **Table S7. Baseline characteristics before the propensity score matching in the population in the sensitivity analysis II, which includes with comorbid cardiovascular disease**

| **Variables** | **AS/WW**  **(n = 1,358)** | **PT**  **(n = 7,180)** | **RT**  **(n = 534)** | **SMD** |
| --- | --- | --- | --- | --- |
| Age at PC diagnosis (mean (SD)) | 62.8 (4.6) | 62.8 (4.6) | 63.2 (4.7) | 0.067 |
| Income levels |  |  |  | 0.137 |
| Low | 265 (19.5) | 1,395 (19.4) | 143 (26.8) |  |
| Middle | 292 (21.5) | 1,565 (21.8) | 104 (19.5) |  |
| High | 801 (59.0) | 4,220 (58.8) | 287 (53.7) |  |
| PC diagnosis calendar year |  |  |  | 0.074 |
| 2012-2013 | 224 (16.5) | 1,669 (23.2) | 132 (24.7) |  |
| 2014-2016 | 269 (19.8) | 1,268 (17.7) | 103 (19.3) |  |
| 2017-2019 | 729 (53.7) | 4,243 (59.1) | 299 (56.0) |  |
| Smoking |  |  |  | 0.052 |
| Never | 490 (36.1) | 2,602 (36.3) | 186 (34.8) |  |
| Former | 630 (46.4) | 3,462 (48.2) | 251 (47.0) |  |
| Current | 238 (17.5) | 1,114 (15.5) | 97 (18.2) |  |
| BMI (kg/m^2^), median (IQR) | 24.1 (2.5) | 24.1 (2.5) | 24.4 (2.6) | 0.081 |
| CCI, mean (SD) | 3.7 (1.4) | 3.8 (1.4) | 3.8 (1.5) | 0.053 |
| Comorbidity |  |  |  |  |
| Hypertension | 672 (49.5) | 3,495 (48.7) | 268 (50.2) | 0.020 |
| Dyslipidemia | 596 (43.9) | 3,112 (43.3) | 235 (44.0) | 0.009 |
| Diabetes mellitus | 389 (28.6) | 2,107 (29.3) | 162 (30.3) | 0.025 |
| BPH | 1,281 (94.3) | 6,945 (96.7) | 520 (97.4) | 0.103 |
| Concomitant drug |  |  |  |  |
| 5-ARI | 256 (18.9) | 1,560 (21.7) | 109 (20.4) | 0.048 |
| Anticholinergic | 102 (7.5) | 666 (9.3) | 46 (8.6) | 0.042 |
| Alpha-blocker | 897 (66.1) | 5,077 (70.7) | 372 (69.7) | 0.067 |
| B3AR | 105 (7.7) | 479 (6.7) | 49 (9.2) | 0.062 |
| Steroid | 753 (55.4) | 4,075 (56.8) | 301 (56.4) | 0.018 |
| Antidepressant | 41 (3.0) | 207 (2.9) | 14 (2.6) | 0.016 |
| AS: active surveillance; WW: watchful waiting; RT: radiotherapy; PT: Prostatectomy; SMD: standardized mean difference; SD: standard deviation; PC: prostate cancer; IQR: interquartile range; CCI: Charlson comorbidity index; BPH: benign prostate hyperplasia; 5-ARI: 5-alpha reductase inhibitor; B3AR: beta 3 adrenoreceptor agonist. | | | | |

### **Table S8. Baseline characteristics after the propensity score matching in the population in the sensitivity analysis II, which includes patients with comorbid cardiovascular disease**

| **Variables** | **AS/WW**  **(n = 1,332)** | **PT**  **(n = 1,332)** | **SMD** | **AS/WW**  **(n = 491)** | **RT**  **(n = 491)** | **SMD** |
| --- | --- | --- | --- | --- | --- | --- |
| Age at PC diagnosis (mean (SD)) | 62.8 (4.7) | 62.9 (4.6) | 0.013 | 63.0 (4.5) | 63.3 (4.6) | 0.060 |
| Income levels |  |  |  |  |  | 0.084 |
| Low | 263 (19.7) | 266 (20.0) | 0.006 | 111 (22.6) | 112 (22.8) |  |
| Middle | 285 (21.4) | 284 (21.3) |  | 104 (21.2) | 103 (21.0) |  |
| High | 784 (58.9) | 782 (58.7) |  | 276 (56.2) | 276 (56.2) |  |
| PC diagnosis calendar year |  |  | 0.025 |  |  | 0.045 |
| 2012-2013 | 223 (16.7) | 226 (17.0) |  | 106 (21.6) | 123 (25.1) |  |
| 2014-2016 | 554 (41.6) | 538 (40.4) |  | 89 (18.1) | 95 (19.3) |  |
| 2017-2019 | 555 (41.7) | 568 (42.6) |  | 287 (58.5) | 273 (55.6) |  |
| Smoking |  |  | 0.036 |  |  | 0.064 |
| Never | 484 (36.3) | 505 (37.9) |  | 185 (37.7) | 170 (34.6) |  |
| Former | 617 (46.3) | 595 (44.7) |  | 222 (45.2) | 233 (47.5) |  |
| Current | 231 (17.3) | 232 (17.4) |  | 84 (17.1) | 88 (17.9) |  |
| BMI (kg/m^2^), mean (SD) | 24.1 (2.5) | 24.0 (2.5) | 0.047 | 24.3 (2.5) | 24.4 (2.6) | 0.041 |
| CCI, mean (SD) | 3.7 (1.4) | 3.8 (1.4) | 0.036 | 3.9 (1.4) | 3.8 (1.4) | 0.060 |
| Comorbidity |  |  |  |  |  |  |
| Hypertension | 658 (49.4) | 636 (47.7) | 0.033 | 258 (52.5) | 247 (50.3) | 0.045 |
| Dyslipidemia | 587 (44.1) | 582 (43.7) | 0.008 | 214 (43.6) | 219 (44.6) | 0.021 |
| Diabetes mellitus | 386 (29.0) | 398 (29.9) | 0.020 | 149 (30.3) | 149 (30.3) | <0.001 |
| BPH | 1,277 (95.9) | 1,271 (95.4) | 0.022 | 483 (98.4) | 482 (98.2) | 0.016 |
| Concomitant drug |  |  |  |  |  |  |
| 5-ARI | 255 (19.1) | 271 (20.3) | 0.030 | 86 (17.5) | 99 (20.2) | 0.068 |
| Anticholinergic | 102 (7.7) | 102 (7.7) | <0.001 | 41 (8.4) | 44 (9.0) | 0.022 |
| Alpha-blocker | 895 (67.2) | 899 (67.5) | 0.006 | 334 (68.0) | 345 (70.3) | 0.049 |
| B3AR | 105 (7.9) | 89 (6.7) | 0.046 | 42 (8.6) | 47 (9.6) | 0.035 |
| Steroid | 742 (55.7) | 739 (55.5) | 0.005 | 280 (57.0) | 279 (56.8) | 0.004 |
| Antidepressant | 39 (2.9) | 47 (3.5) | 0.034 | 9 (1.8) | 14 (2.9) | 0.067 |
| AS: active surveillance; WW: watchful waiting; RT: radiotherapy; PT: Prostatectomy; SMD: standardized mean difference; SD: standard deviation; PC: prostate cancer; IQR: interquartile range; CCI: Charlson comorbidity index; BPH: benign prostate hyperplasia; 5-ARI: 5-alpha reductase inhibitor; B3AR: beta 3 adrenoreceptor agonist. | | | | | | |

### **Table S9. Incidence and hazard ratios of secondary outcomes in the sensitivity analysis I**

| **Variables** | **AS/WW** | **PT** | **AS/WW** | **RT** |
| --- | --- | --- | --- | --- |
|  | **(N=2,239)** | **(N=2,239)** | **(N=1,160)** | **(N=1,160)** |
| Primary outcome |  |  |  |  |
| Prostate cancer death |  |  |  |  |
| No. of events | 19 | 8 | 13 | 16 |
| PY | 9,661 | 9,805 | 5,128 | 5,072 |
| IR [95% CI] | 1.97 [1.18-3.07] | 0.82 [0.35-1.61] | 2.54 [1.35-4.33] | 3.15 [1.80-5.12] |
| HR [95% CI] | Reference | 0.42 [0.18-0.95] | Reference | 1.25 [0.60-2.59] |
| Secondary outcomes |  |  |  |  |
| Death from any cause |  |  |  |  |
| No. of events | 83 | 66 | 57 | 54 |
| No. of PY | 9,661 | 9,805 | 5,128 | 5,072 |
| IR [95% CI] | 8.59 [6.84-10.70] | 6.73 [5.21-8.56] | 11.1 [8.42-14.4] | 10.6 [8.00-13.9] |
| HR [95% CI] | Reference | 0.78 [0.56-1.08] | Reference | 0.96 [0.66-1.40] |
| Metastatic disease |  |  |  |  |
| No. of events | 275 | 220 | 173 | 160 |
| PY | 8857 | 9521 | 4634 | 4687 |
| IR [95% CI] | 31.0 [27.5-34.9] | 23.8 [20.7-27.1] | 37.3 [32.0-43.3] | 34.1 [29.1-39.9] |
| HR [95% CI] | Reference | 0.77 [0.65-0.92] | Reference | 0.91 [0.74-1.13] |
| Initiation of antidepressant | |  |  |  |
| No. of events | 317 | 309 | 168 | 115 |
| PY | 8743 | 8927 | 4675 | 4755 |
| IR [95% CI] | 36.3 [32.4-40.5] | 34.6 [30.9-38.7] | 34.9 [29.7-40.7] | 24.2 [20.0-29.0] |
| HR [95% CI] | Reference | 0.96 [0.82-1.12] | Reference | 0.69 [0.54-0.89] |

AS/WW: active surveillance/watchful waiting; PT: prostatectomy; RT: radiotherapy; No: number; PY: person-year; IR: incidence rate per 1,000 person-year; HR: hazard ratio; CI: confidence interval.

### **Table S10. Incidence and hazard ratios of secondary outcomes in the sensitivity analysis II**

| **Variables** | **AS/WW** | **PT** | **AS/WW** | **RT** |
| --- | --- | --- | --- | --- |
|  | **(N=1,332)** | **(N=1,332)** | **(N=491)** | **(N=491)** |
| Primary outcome |  |  |  |  |
| Prostate cancer death | |  |  |  |
| No. of events | 2 | 2 | 0 | 4 |
| PY | 5840 | 5839 | 2254 | 2202 |
| IR [95% CI] | 0.34 [0.04-1.24] | 0.34 [0.04-1.24] | NA | 1.82 [0.49-4.65] |
| HR [95% CI] | Reference | 1.01 [0.14-7.16] | Reference | NA |
| Secondary outcomes | |  |  |  |
| Death from any cause | |  |  |  |
| No. of events | 19 | 14 | 5 | 12 |
| PY | 5840 | 5839 | 2254 | 2202 |
| IR [95% CI] | 3.25 [1.96-5.08] | 2.40 [1.31-4.02] | 2.22 [0.72-5.18] | 5.45 [2.82-9.52] |
| HR [95% CI] | Reference | 1.35 [0.37-1.48] | Reference | 2.46 [0.87-6.98] |
| Metastatic disease |  |  |  |  |
| No. of events | 138 | 110 | 50 | 50 |
| PY | 5439 | 5569 | 2118 | 2065 |
| IR [95% CI] | 25.4 [21.3-30.0] | 19.8 [16.2-23.8] | 23.6 [17.5-31.1] | 24.2 [18.0-31.9] |
| HR [95% CI] | Reference | 0.78 [0.61-1.00] | Reference | 1.02 [0.69-1.52] |
| Initiation of antidepressant | |  |  |  |
| No. of events | 180 | 148 | 69 | 37 |
| PY | 5300 | 5421 | 2023 | 2087 |
| IR [95% CI] | 34.0 [29.2-39.3] | 27.3 [23.1-32.1] | 34.1 [26.5-43.2] | 17.7 [12.5-24.4] |
| HR [95% CI] | Reference | 0.81 [0.65-1.01] | Reference | 0.52 [0.35-0.78] |

AS/WW: active surveillance/watchful waiting; PT: prostatectomy; RT: radiotherapy; No: number; PY: person-year; IR: incidence rate; HR: hazard ratio; CI: confidence interval.

### **Table S11. Hazard ratios of prostate specific mortality in the competing risk analysis**

| **Variables** | **Hazard ratio [95% CI]** | | |
| --- | --- | --- | --- |
|  | **AS/WW** | **PT** | **RT** |
| Prostate cancer death |  |  |  |
| Main setting | Ref | 1.01 [0.14-7.19] | 1.72 [0.56-5.30] |
| Sensitivity I: Age >40 | Ref | 0.42 [0.18-0.95] | 1.12 [0.77-1.61] |
| Sensitivity II: Population including comorbid CVD | Ref | 1.01 [0.14-7.08] | NA |
| CI: confidence interval; AS/WW: active surveillance/watchful waiting; PT: prostatectomy; RT: radiotherapy; CVD: cardiovascular disease; Ref: reference; NA: not available. | | | |

### **Figure S1. Schematic diagram for the cohort definition in the main setting**

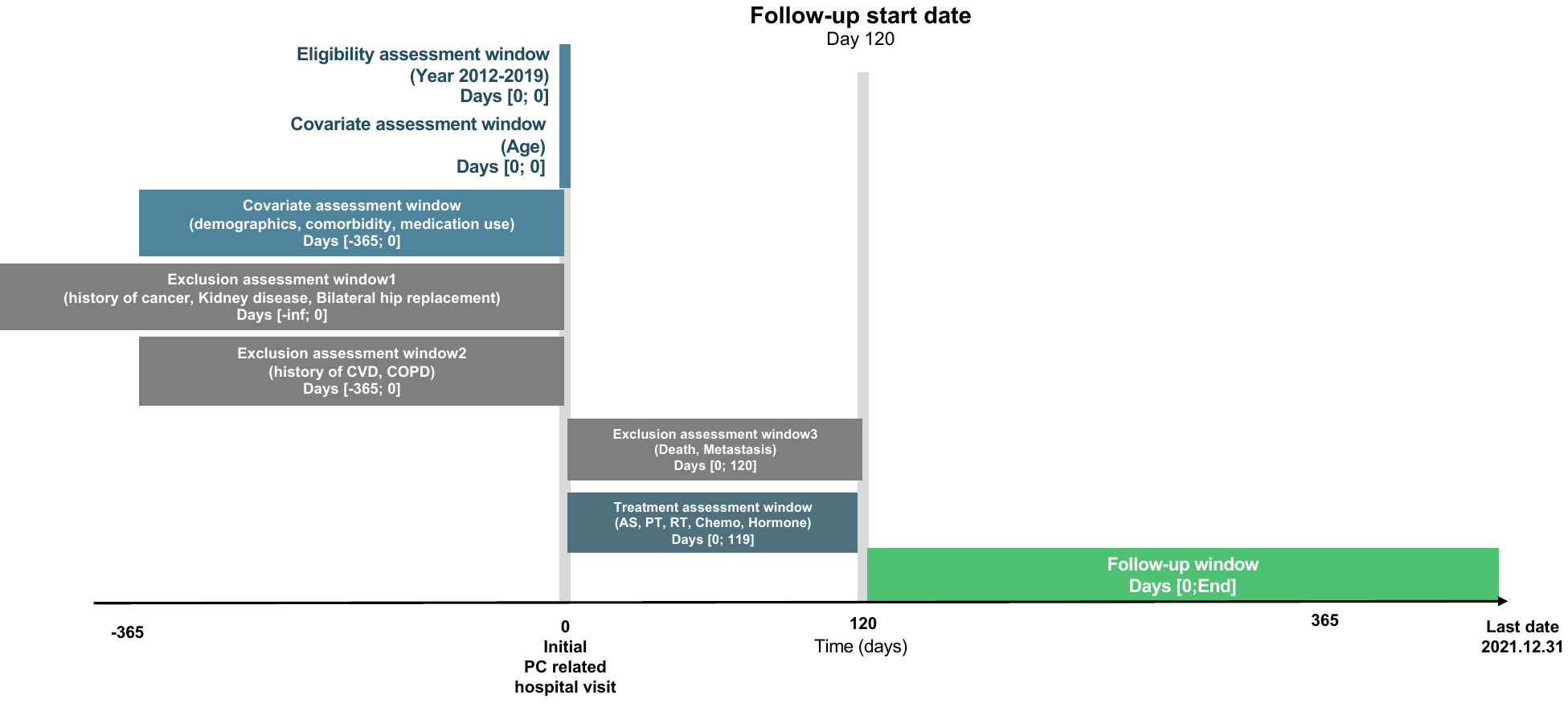

### **Figure S2. Cumulative incidence plots for metastasis and antidepressant initiation when comparing PT to AS/WW in the main setting**

(a) Metastasis	(b) Antidepressant initiation

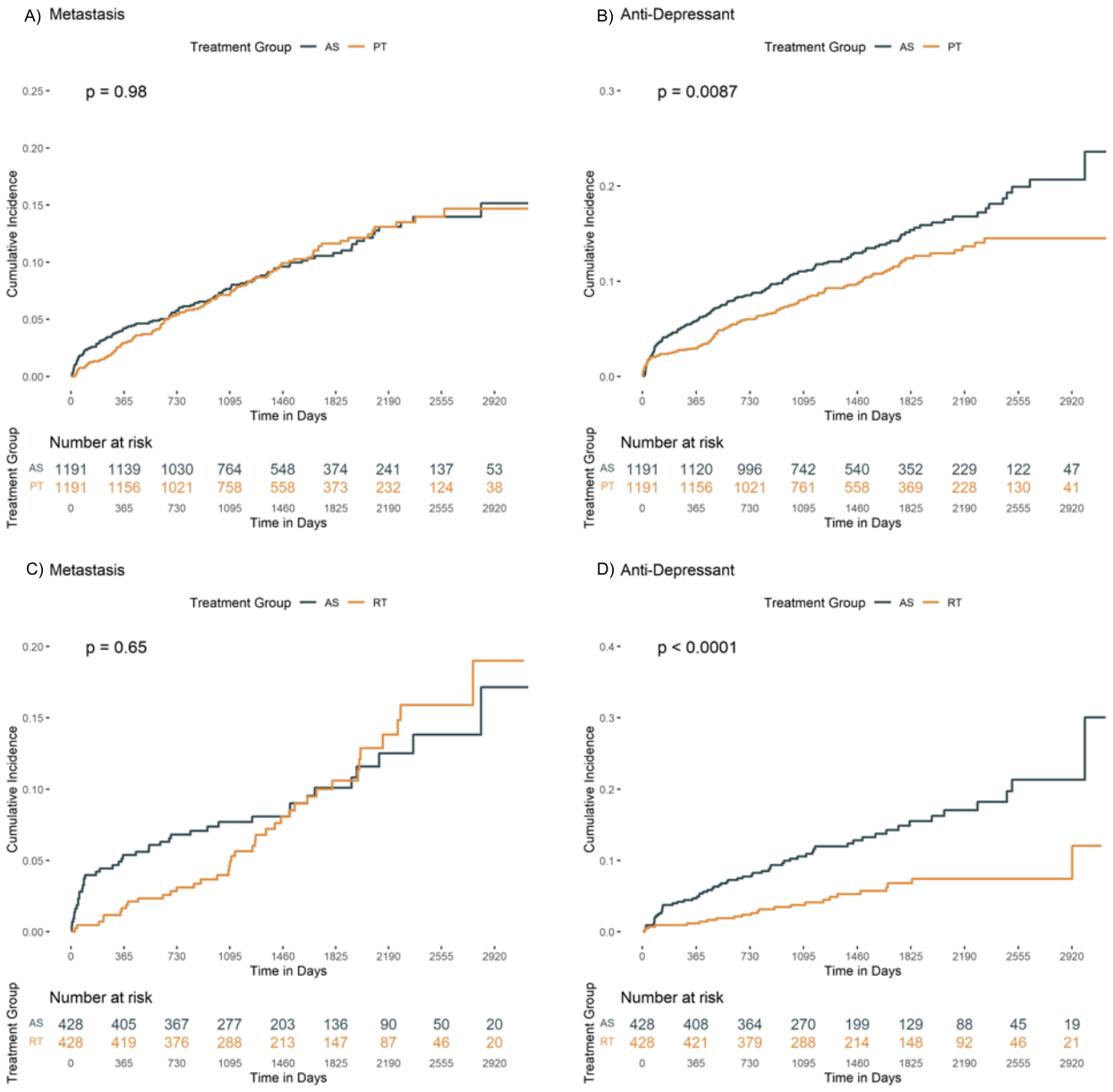

PT: prostatectomy; AS/WW: active surveillance/watchful waiting

### **Figure S3. Cumulative incidence plots for metastasis and antidepressant initiation when comparing RT to AS/WW in the main setting**

(a) Metastasis	(b) Antidepressant initiation

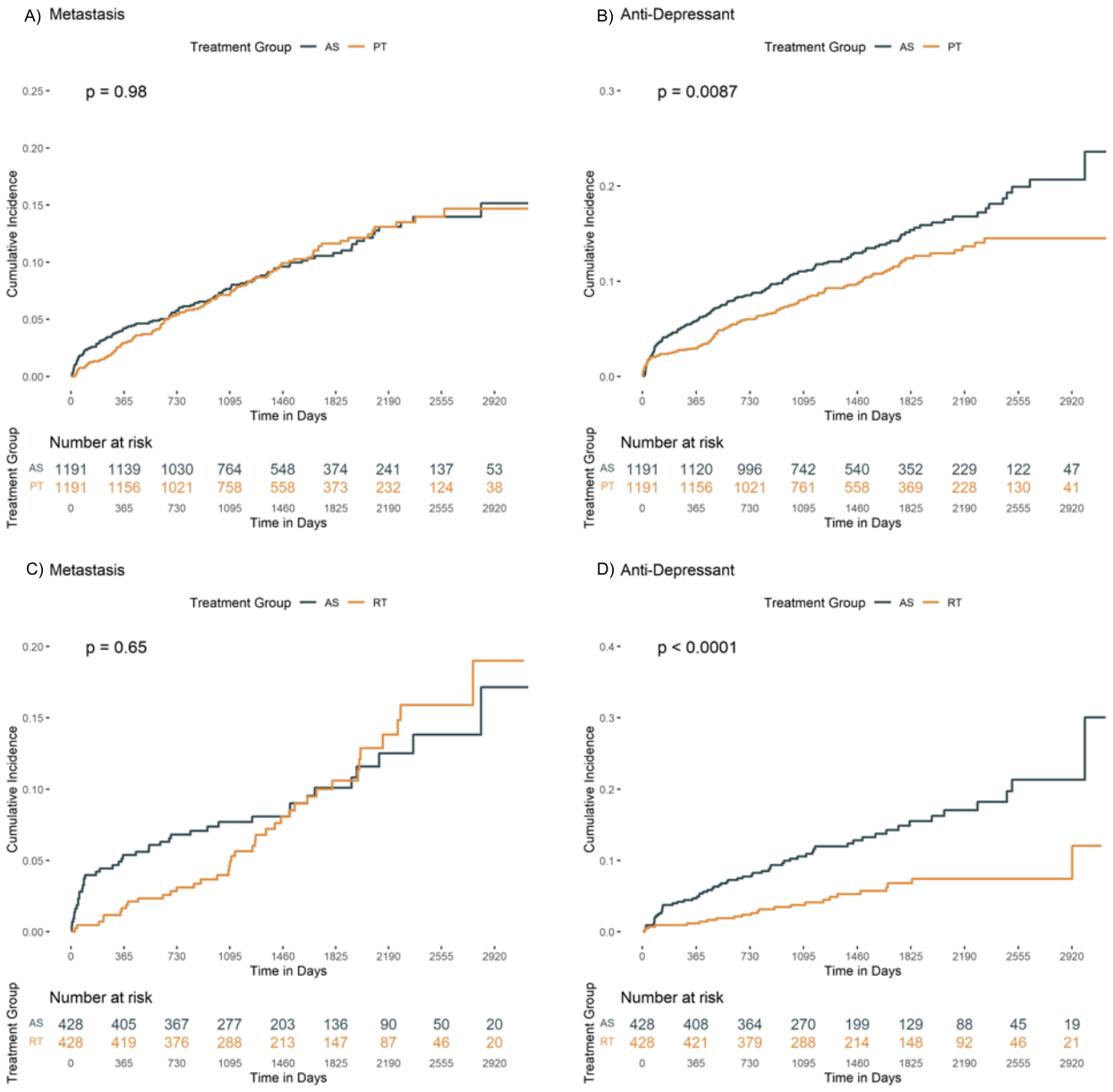

PT: prostatectomy; AS/WW: active surveillance/watchful waiting

### **Figure S4. Cumulative incidence plots for outcomes comparing PT vs AS/WW in the sensitivity analysis I (age over 40 years old)**

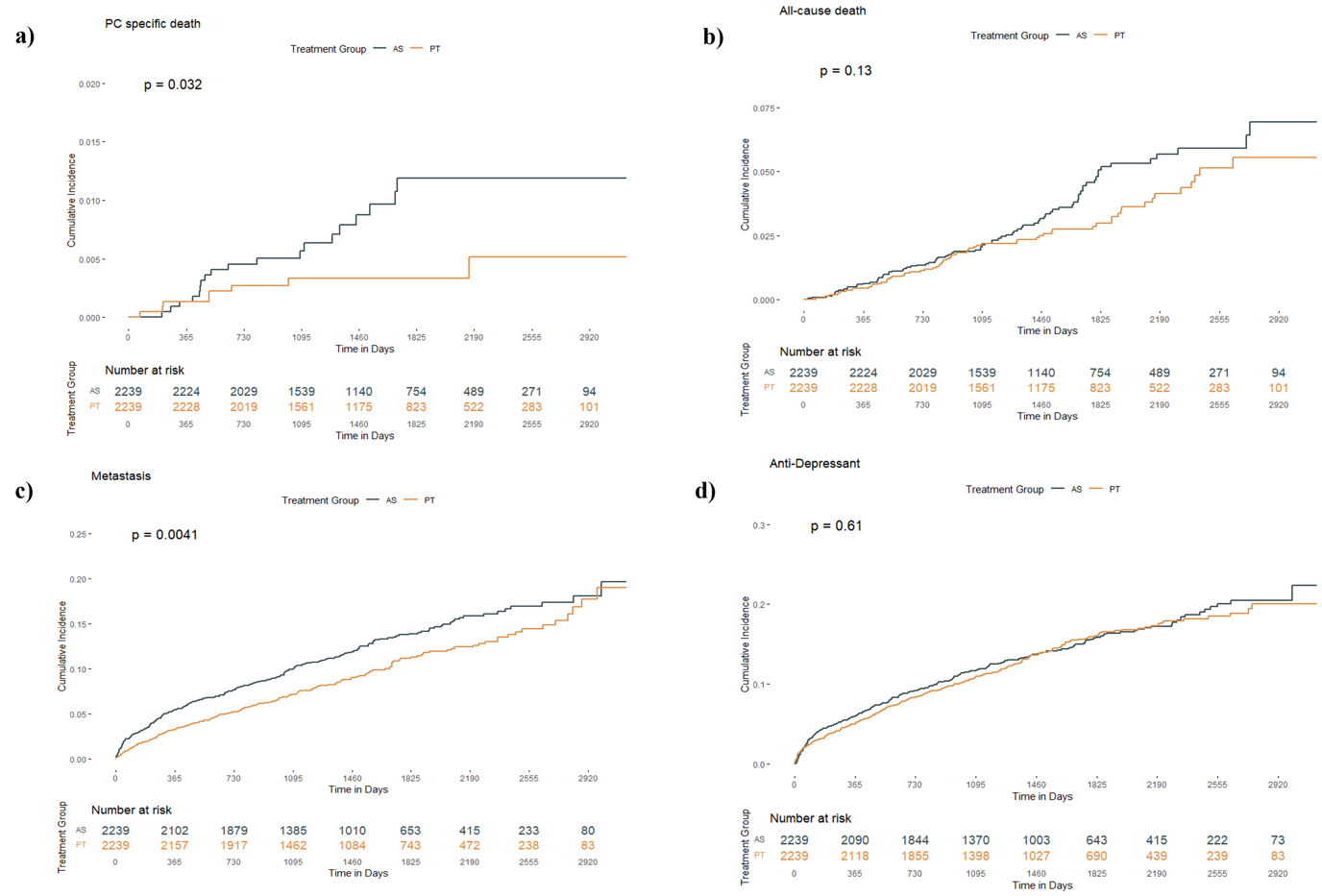

a) prostate-specific death, b) all-cause death, c) metastasis, and d) antidepressant initiation. PT: prostatectomy; AS/WW: active surveillance/watchful waiting.

### **Figure S5. Cumulative incidence plots for outcomes comparing RT vs AS/WW in the sensitivity analysis I (age > 40 years old)**

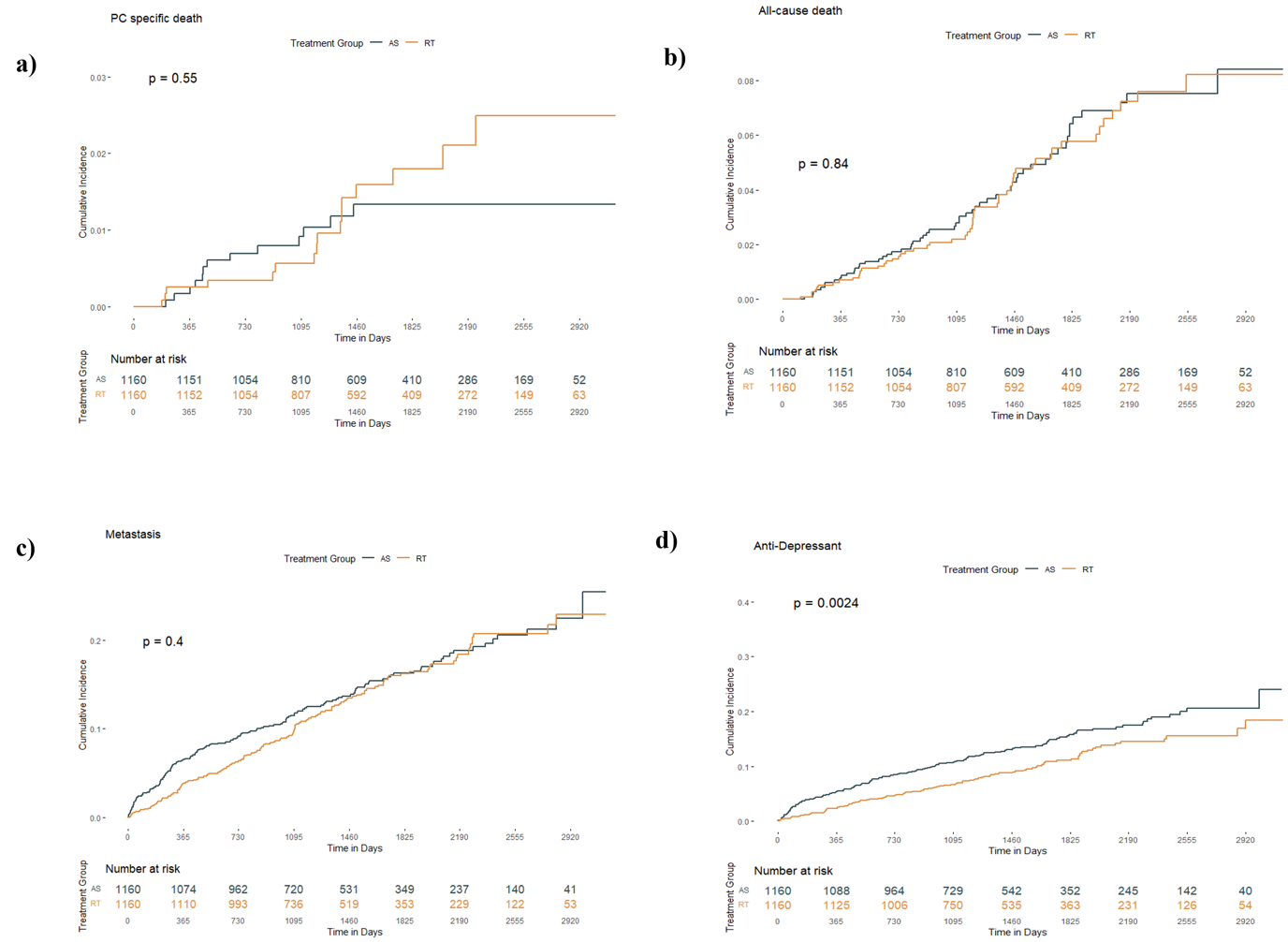

a) prostate-specific death, b) all-cause death, c) metastasis, and d) antidepressant initiation. RT: radiotherapy; AS/WW: active surveillance/watchful waiting.

### **Figure S6. Cumulative incidence plots for outcomes comparing PT vs AS/WW in the sensitivity analysis II (including patients with cardiovascular comorbidities)**

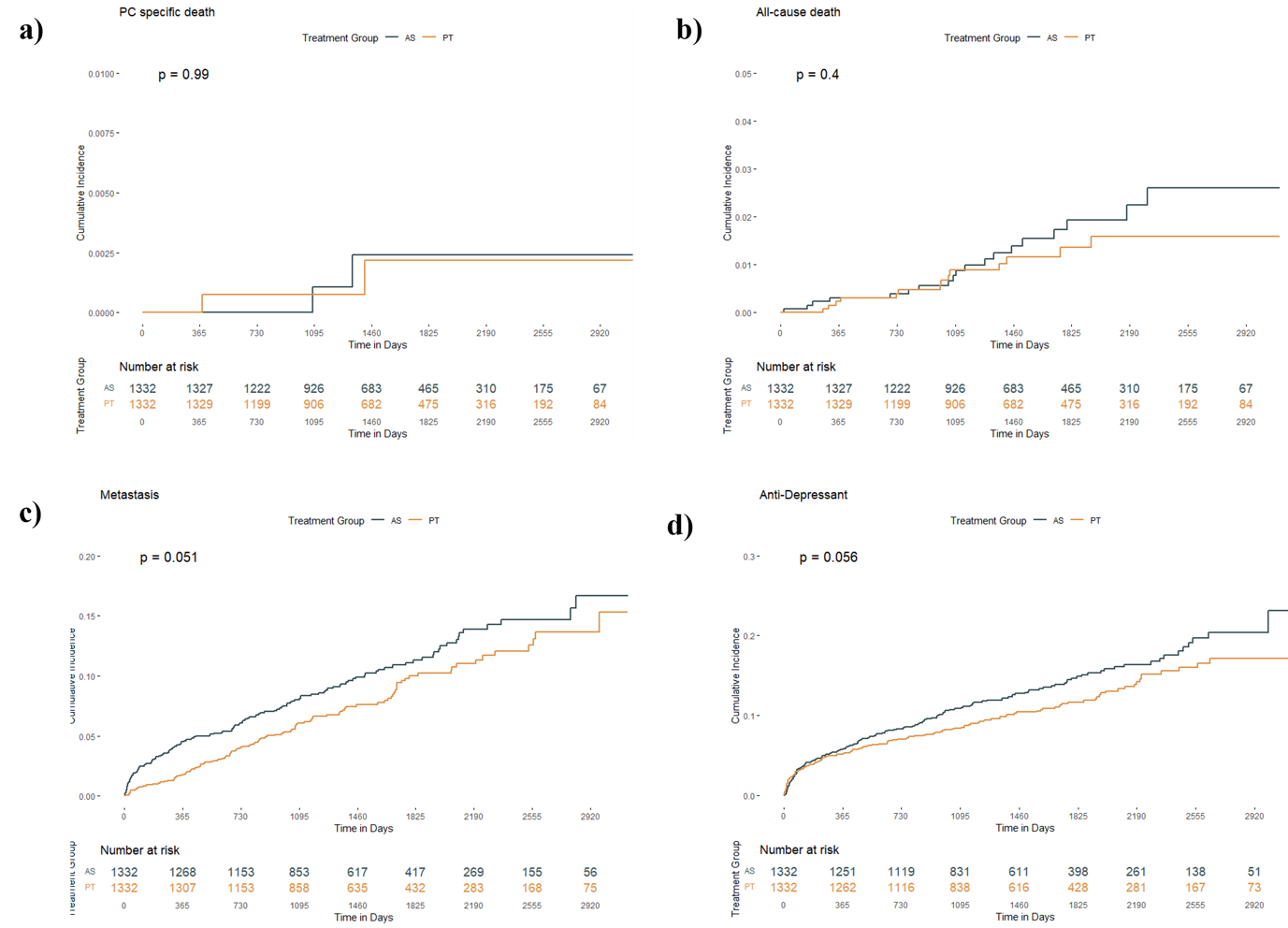

a) prostate-specific death, b) all-cause death, c) metastasis, and d) antidepressant initiation. PT: prostatectomy; AS/WW: active surveillance/watchful waiting.

### **Figure S7. Cumulative incidence plots for outcomes comparing RT vs AS/WW in the sensitivity analysis II (including patients with cardiovascular disease)**

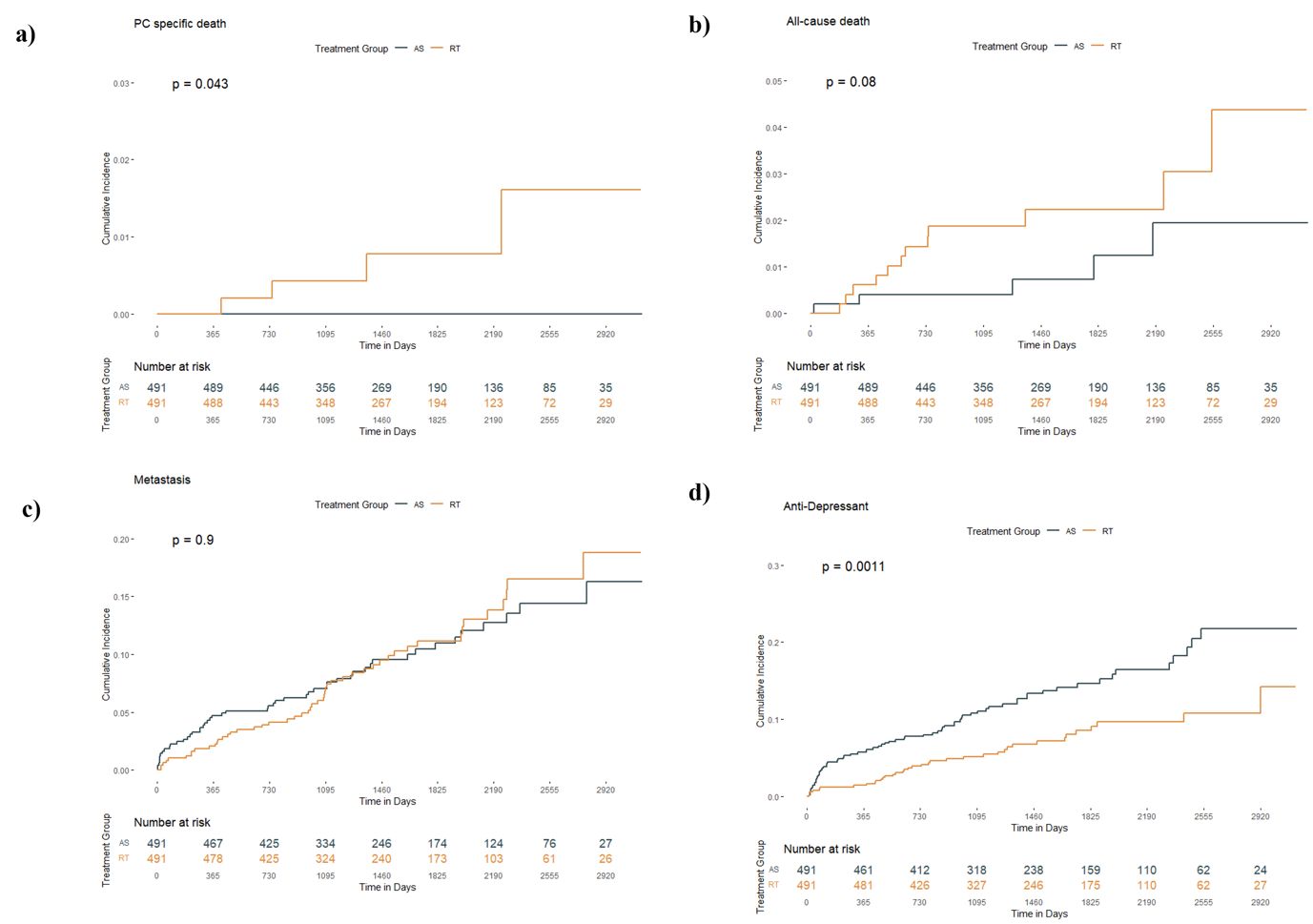

a) prostate-specific death, b) all-cause death, c) metastasis, and d) antidepressant initiation. RT: radiotherapy; AS/WW: active surveillance/watchful waiting.
